# Early epidemiological signatures of novel SARS-CoV-2 variants: establishment of B.1.617.2 in England

**DOI:** 10.1101/2021.06.05.21258365

**Authors:** Robert Challen, Louise Dyson, Christopher E. Overton, Laura M. Guzman-Rincon, Edward M. Hill, Helena B. Stage, Ellen Brooks-Pollock, Lorenzo Pellis, Francesca Scarabel, David J. Pascall, Paula Blomquist, Michael Tildesley, Daniel Williamson, Stefan Siegert, Xiaoyu Xiong, Ben Youngman, Juniper, Jonathan M. Read, Julia R. Gog, Matthew J. Keeling, Leon Danon

## Abstract

The rapid emergence of SARS-CoV-2 mutants with new phenotypic properties is a critical challenge to the control of the ongoing pandemic. B.1.1.7 was monitored in the UK through routine testing and S-gene target failures (SGTF), comprising over 90% of cases by March 2021. Now, the reverse is occurring: SGTF cases are being replaced by an S-gene positive variant, which we associate with B.1.617.2. Evidence from the characteristics of S-gene positive cases demonstrates that, following importation, B.1.617.2 is transmitted locally, growing at a rate higher than B.1.1.7 and a doubling time between 5-14 days. S-gene positive cases should be prioritised for sequencing and aggressive control in any countries in which this variant is newly detected.

**One-Sentence Summary:** The B.1.617.2 variant of SARS-CoV-2 is replacing B.1.1.7 and emerging as the dominant variant in England, evidenced by sustained local transmission.

---

SARS-CoV-2 has caused millions of cases and deaths worldwide, generating thousands of variants that are circulating globally (*1*). Most mutations show no detectable phenotypic change and no selective advantage, however other mutations have emerged that confer higher transmission or vaccine escape potential (*2*). These are termed “variants-of-concern” (VOCs) and pose a serious threat to disease control.

As of May 2021, current VOCs include B.1.1.7, which emerged in southeast England in September 2020. Its large number of accrued mutations conferred increased transmissibility compared to earlier variants (*3, 4*), and increased mortality (*5, 6*). B.1.351, which emerged in South Africa, shares some of the same mutations as B.1.1.7, but also appears to show reduced sensitivity to immune responses acquired against the ‘wild-type’ Wuhan virus (*7*) or generated by current vaccines (*8*). B.1.617, the lineage behind the large number of cases in India in 2021, is currently increasing as a proportion of sequenced lineages in the UK against a background of B.1.1.7 sequences, suggesting a competitive advantage. B.1.617 comprises three sublineages: B.1.617.1, B.1.617.2 and B.1.617.3 (*9*). Two of the sublineages, B.1.617.1 and B.1.617.3 have the E484Q mutation, which may reduce viral neutralisation and could facilitate vaccine escape (*10*), and have been designated as “variants under investigation” (VUIs). For B.1.617.2, designated as a VOC on 6 May 2021 (*11*), there is emerging evidence on sensitivity to vaccine-acquired immunity with early studies suggesting no clear change (*12*) and experimental studies showing reduced sensitivity (*13*). Variants of SARS-CoV-2 that show increased transmissibility or escape from vaccine-derived immunity, even partially, could generate large future waves of infection requiring further costly NPIs to prevent healthcare systems being overwhelmed (*14*). Early identification and control of VOCs is therefore essential for limiting their impact.

Here, we present multiple lines of evidence indicating the variant of concern B.1.617.2 is in the early stages of invasion in England. We combine genetic information from sequencing data with S-gene positivity status from testing data (which identifies B.1.617.2 and other variants but not the dominant B.1.1.7), and link this to demographic and geographical data to investigate the recent increase in cases and associate it with the local establishment of the variant of concern, B.1.617.2.

## Identifying variants in routine UK surveillance data

The emergence and progression of the variant B.1.1.7 in the UK could be observed because, in contrast to previously circulating ‘wild-type’ variants, B.1.1.7 exhibits a deletion in the SARS-CoV-2 genome at site 69-70 associated with the spike protein, leading to the ThermoFisher TaqPath quantitative PCR assay failing to amplify the S-gene target (*15*). The percentage of cases with an S-gene target failure, or SGTF, increased from 3% in October 2020 to 98% by December 2020 (*16*). In contrast, other designated variants-of-concern including B.1.351 (first described in South Africa), P.1 (first described in Brazil) and B.1.617 lineages (first described in India, a sublineage of which B.1.617.2 was designated a VOC in April 2021) are all S-gene positive on the TaqPath assay (*15*) as they do not have the same spike protein deletion.

The high prevalence of B.1.1.7 and associated S-gene negatives (*17*) allows us to use S-gene positive cases from the TaqPath assay in community testing (known as Pillar 2 in the UK) as a rapid signal for investigating community spread of S-gene positive variants, including B.1.351 and B.1.617. Early 2021 was characterised by falling S-gene positive and S-gene negative cases, but since mid-March some regions have seen an increase in the relative proportion of S-gene positive cases (Figure 1A). Since early April 2021 there has been a steady rise in the absolute number of S-gene positive cases in some regions while S-gene negative cases continue to fall. S-gene positive cases are overtaking S-gene negative cases at the time of writing (start of June 2021).

**Figure 1.**
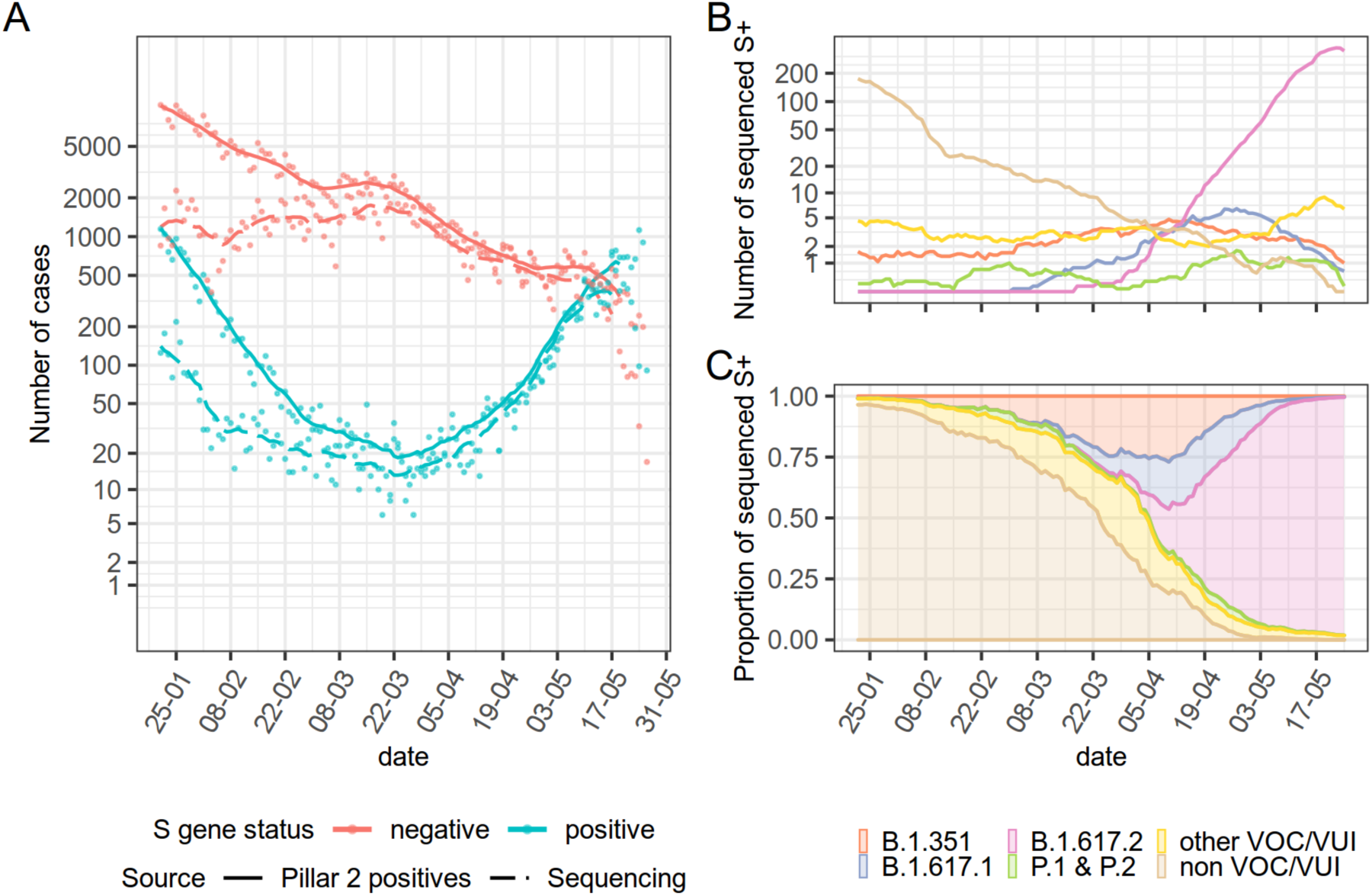
Temporal patterns in SARS-CoV-2 variants in England: (A) Pillar 2 S-gene positive and negative case counts (solid lines) since November 2020 (14-day rolling average) and sequencing activity associated with those cases (dashed lines). S-gene positive cases have been growing exponentially since late March. (B) The number of cases of different variants, from sequencing samples of S-gene positive tests (14-day rolling average). (C) Proportion of S-gene positive cases of different variants, showing recent dominance of B.1.617.2 over other VOC/VUI (14-day rolling average). Delays in returning sequencing results compared to S-gene tests lead to an underestimate of the expected count of B.1.617.2 (panel B) at the most recent time point.

In contrast to sequencing, which has a reporting delay of up to 3 weeks, the presence or absence of an S-gene from the TaqPath assay is obtained with shorter delay and no additional extra resources. Examining S-gene positive cases that have been sequenced in more detail reveals that the variant responsible for the increase in S-gene positive case numbers is B.1.617.2, the current dominant variant in India (Fig. 1B). Over the time period of interest, all other S-positive variants have either remained stable or declined. From 1st May 2021, B.1.617.2 is the proven cause of more than 95% of all S-positive cases, when sequenced (Fig. 1C). The association between S-gene positive cases and B.1.617.2 is supported by comparison of their geographic distributions in Supplementary Materials Section 5.

## Results

### Relative growth rates of B.1.617.2 and B.1.1.7

We measured the growth rate of the S-gene positive and S-gene negative cases to assess the relative competitive advantage of VOCs, given the increasingly close relationship between B.1.617.2 and S- gene positive cases, and the well-established relationships between S-gene negative cases and B.1.1.7. The growth rate describes the exponential rate at which cases are growing or declining in a given area. As opposed to the widely used reproduction number (*18*), the growth rate can be estimated directly from data and provides a reliable measure of the speed of growth of cases regardless of whether they derive from direct transmission or other sources such as importation. We estimated the instantaneous growth rate using four independent methods, each based on different assumptions and modelling techniques: a Generalised Additive Model, a Gaussian Process model, a Poisson regression, and a Poisson model fitted in a Bayesian framework (see Supplementary Materials Section 3 for additional details).

In England, the estimated instantaneous growth rate of S-gene negative cases appears relatively stable in the observed time window, from the beginning of February to mid-May 2021, indicating that cases were consistently declining (Figure 2A). Conversely, a clear increase in S-gene positive cases since the beginning of April is confirmed (Figure 1A), with growth rate estimates remaining above zero (Figure 2A) with doubling times as short as 7 days in early May 2021. The transition from comparatively lower growth rates of S-gene positive cases at the beginning of the time series, to comparatively higher growth rates of S-gene positive cases at the end of the time series mirrors the transition seen in Figure 1B from primarily “non-VOC/VUI” cases at the beginning to B.1.617.2 cases towards the end.

**Figure 2.**
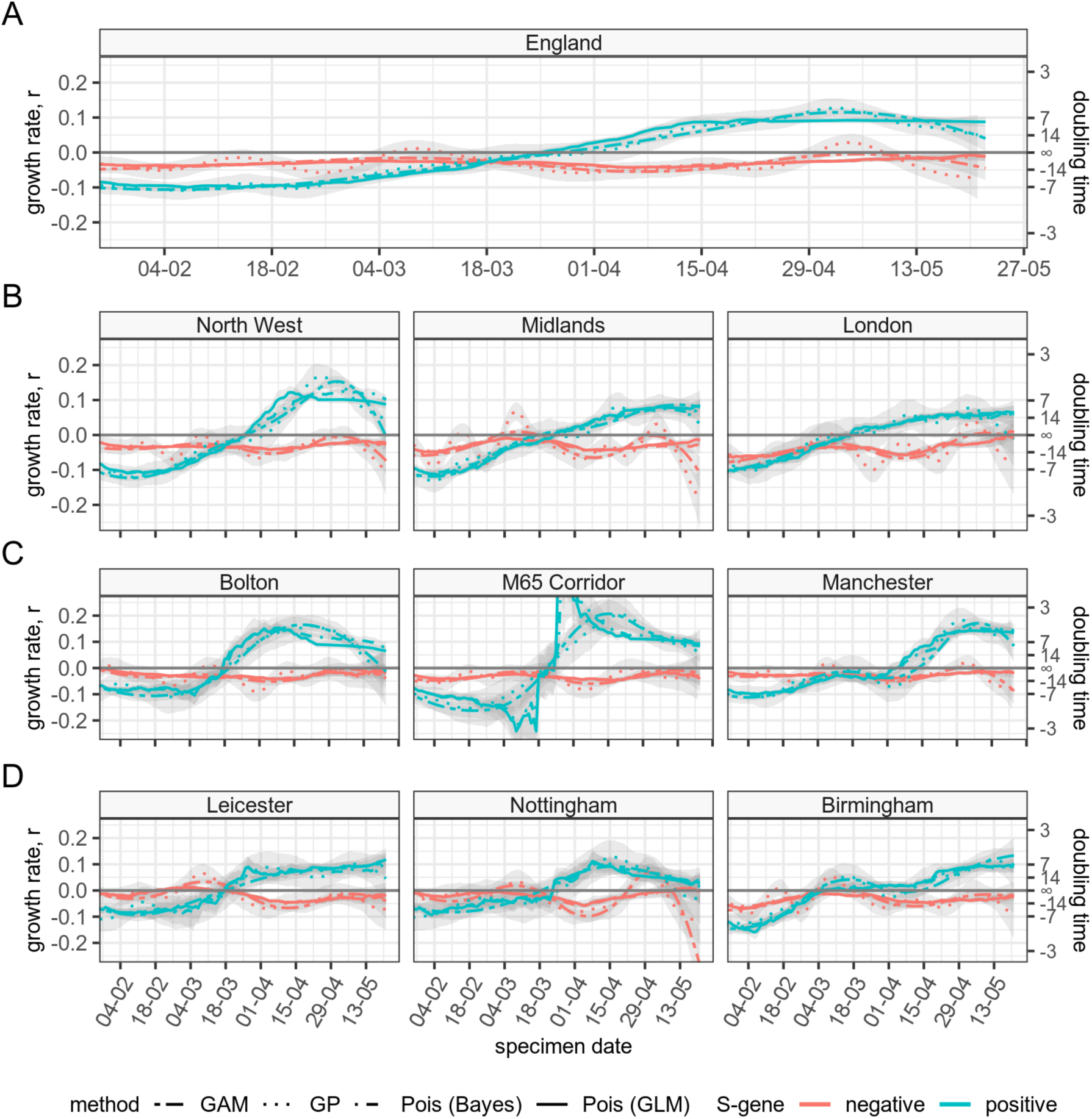
Estimated growth rate and doubling time for S-gene positive and S-gene negative symptomatic cases in England. Green lines denote S-gene positive cases and red denote S-gene negative cases. Four methods used to estimate growth rate time series are denoted with full line (Poisson regression method), dotted line (Gaussian Process method), dashed line (Generalised Additive Model method), and dot-dash (Poisson Bayesian method). Panels show a hierarchy of areas: (A) England, (B) NHS Regions which are regions within (A), (C) Cities and sub-regions of the North-West, (D) Cities in the Midlands. The M65 corridor includes the towns of Blackburn, Accrington and Burnley. See Supplementary Materials for precise geographic definitions and details of growth rate estimation methods.

Spatially aggregated data may smooth out and hide local trends when epidemics are geographically heterogeneous, so we estimate the instantaneous growth rate for smaller geographies. Figure 2 also shows a selection of regions of England (North West, Midlands, London, Figure 2B) and cities and sub-regions within the North West and Midlands (Figures 2C,D), where the highest growth rate of S-gene positive cases was observed. We estimated a positive growth rate of S-gene positive cases starting from the second half of March consistently across these regions, although with slightly different onsets. The growth of S-gene positive cases appears sustained until the end of the studied period despite a background of generally declining S-gene negative cases (except for Nottingham, showing an increase in both S-gene positive *and* negative cases in May 2021). This suggests a competitive advantage of S- gene positive cases versus S-gene negative. The transition from low growth, “non-VOC/VUI” S-gene positive cases to B.1.617.2 S-gene positive cases in smaller geographic units is more abrupt.

The relative growth rate estimates assume that testing efforts are similar over time. Since testing frequency is only partially observed, it is not possible to adjust for this entirely, so we present a wider set of measures to inform our conclusions; the results from all three methods give very similar results, increasing confidence in the observed patterns. The relative growth rate of S-gene positive compared to S-gene negative cases provides an assessment of their relative competitive advantage. While Figure 2 shows estimates obtained using data from symptomatic cases only, which are likely less affected by testing biases, additional analyses for a different dataset including screening cases are included in the Supplementary Materials (SM Section 3), showing similar conclusions.

### Shifting age distributions of cases

The age distribution of confirmed cases in a generalised epidemic reflects the population mixing patterns and age-stratified infectivity, susceptibility, severity and testing-seeking behaviour. If most B.1.617.2 cases are imported through travel, we would expect the observed age distributions to reflect the age distribution of travellers (see Supplementary Materials Section 3, Figure S3.6). Therefore, prior to importation of B.1.617.2, S-gene positive cases are expected to correspond to other variants in the community, under the assumption that variants do not differ substantially in their age specific susceptibility or infectivity and follow the same age distribution as S-gene negative cases, since both are established. During the time of peak imports, we expect the S-gene positive samples to reflect the age of travellers and their immediate contacts before returning to a distribution reflecting community transmission. We would then expect the age distribution to revert to that of S-gene negative cases.

The dynamics of perturbation and equilibration are summarised with the Wasserstein (or Kantorovich) distance metric (*19, 20*), which captures discrepancies between the age distribution of S-gene negative and positive samples, in Figure 3; findings are confirmed with the Kolmogorov-Smirnov distance in Supplementary Materials Section 3. These metrics are sensitive to sample size, so to assess uncertainty we generate a 95% significance level confidence region for the null hypothesis that S-gene positive and S-gene negative have the same age distribution, based on a permutation test. If the distance metric falls outside this confidence region, the null hypothesis is rejected, and we conclude that the age distributions are different for S-gene positive and S-gene negative cases.

**Figure 3.**
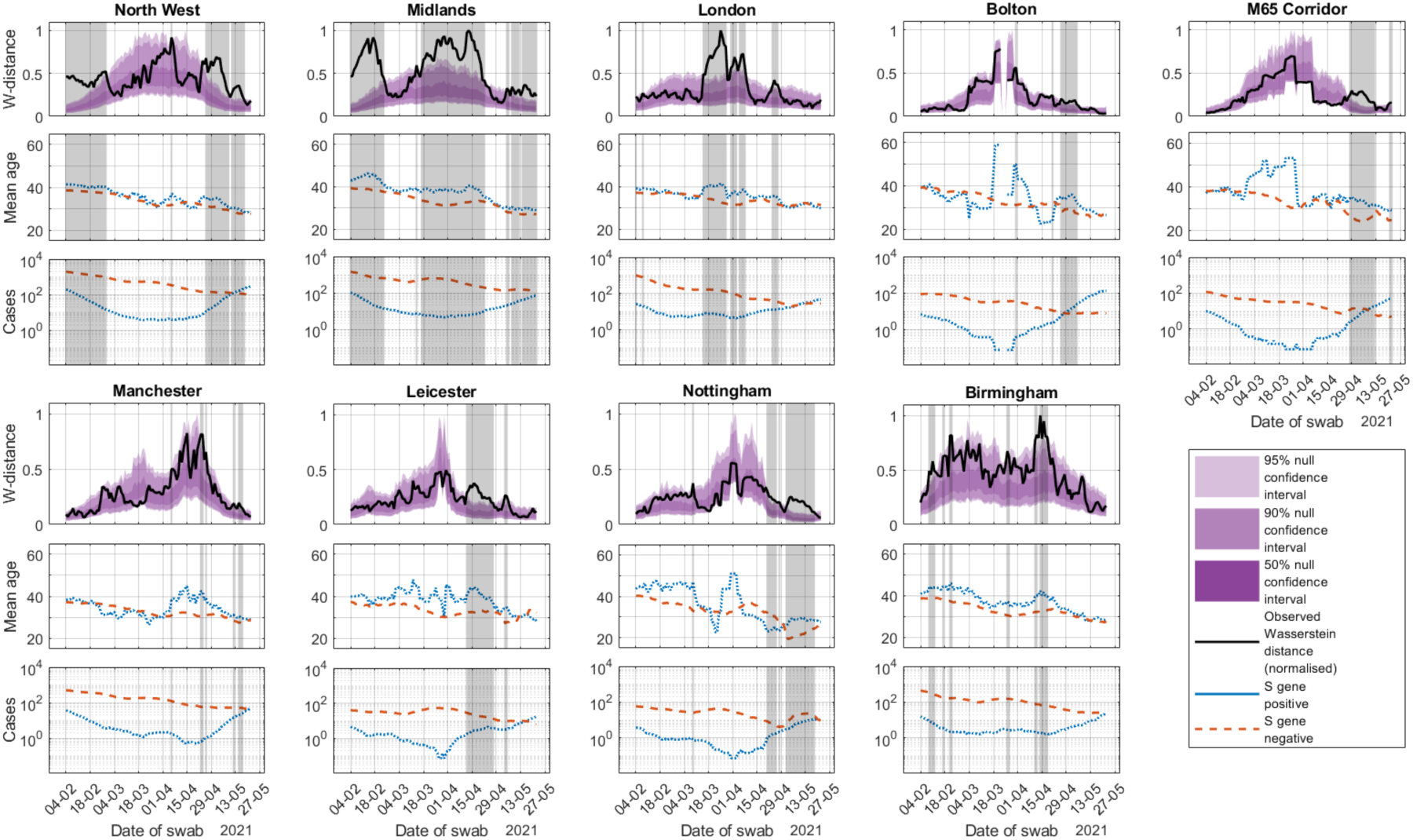
Dynamics of age distributions of cases. For each region, we show the Wasserstein distance between the age distribution of S-gene positive and S-gene negative cases, normalised with peak value at 1 (top panels), the mean age (middle panels) and the number of cases (bottom panels). All metrics are computed over a 14-day rolling window of cases by specimen date, plotted by the last swab date in the 14-day window. The black curve indicates the Wasserstein distance between the two age distributions. The purple shaded region indicates the confidence region for the Wasserstein distance if the two samples were drawn from the same distribution, generated through 1000 Monte Carlo samples of a permutation test (see text and Supplementary Materials Section 3). The grey shaded bands indicate dates where the Wasserstein distance falls outside of the 95% confidence region, denoting a significant difference between the age distribution of S-gene positive and S-gene negative cases. The red dashed curve indicates S-gene negative cases and the blue dotted curve indicates S-gene positive cases.

In Bolton, Leicester and Birmingham the Wasserstein distance rises for short periods of time in late March and early April 2021, leaving the null hypothesis confidence region (Figure 3). This is consistent with the timing and opportunity of importation of B.1.617.2 (see also Supplementary Materials Section 7 for traveller status breakdown). However, on 23 April, travel restrictions were imposed from India, with only British citizens and permanent residents allowed to enter the country subject to a 10-day stay in a quarantine hotel and two mandatory PCR tests (*21*). This rise in the Wasserstein metric followed by its subsequent decline supports the hypothesis that introductions of S-gene positive infections related to B.1.617.2 were quickly followed by sustained transmission in the community, which is reflected in similar age distributions of S-gene positive and S-gene negative cases in the most recent data (Supplementary Materials Section 3). The slightly older average age of cases that are S-gene positive may reflect slightly lower vaccine efficacy against B.1.617.2, such that older age groups who are more likely to have received the vaccine are at elevated risk of B.1.617.2 infection compared to B.1.1.7.

## Discussion

We provide evidence that S-gene positive SARS-CoV-2 infections in England, although initially confined to travellers and their close contacts, have now become established in the wider population. All indications are that these cases are now due to B.1.617.2. Recently, S-gene positive cases have a substantially larger growth rate than S-gene negative cases, which suggests that B.1.617.2 is likely to become the dominant variant in the UK, replacing B.1.1.7. This analysis was only possible by combining genomically incomplete, yet widely collected routine testing data with the delayed signal from genetically complete cases. In early 2021, the majority of positive SARS-CoV-2 tests in England were genomically sequenced (74% in April 2021), with the sequencing subject to processing delays: 21% of sequencing results were available within 1 week, 44% within 2 weeks, and 66% within 3 weeks (as of 27th May 2021). At the end of the study period, nearly all S-gene positive samples that were sent for sequencing were subsequently verified to be B.1.617.2, yet many S-gene positive samples were still being processed. We can draw conclusions about the spread of B.1.617.2 from S-gene data because of the close relationship between S-gene positivity and B.1.617.2 in England at the end of the study period.

The shortest COVID-19 doubling times estimated in England were in March 2020 when SARS-CoV-2 infections were doubling every 3 days (*22*). Here, S-gene positive cases were estimated to be doubling every 7 days in England overall, and as rapidly as every 5 days in some regions with known outbreaks of B.1.617.2. This growth rate is in contrast to a shrinking epidemic of S-gene negative cases in the same areas, and at the same times. Mathematical modelling of the UK’s proposed roadmap to relax social distancing restrictions entirely has demonstrated that variants with even a moderate increase in transmission rate or that partially evade vaccine-derived immunity have the potential to threaten the current (30 May 2021) decline in cases and deaths. Variants with a large competitive advantage can generate a resurgence in cases and hospital admissions larger than experienced in January 2021 in the UK. The rapid doubling of S-gene positive cases calls for speedy and focussed control measures.

In other European countries, a similar picture is emerging. Denmark, Germany and the Netherlands, are seeing B.1.617.2 growing on a background of predominantly B.1.1.7 infections (over 90% of cases in each country). In Denmark, where up to 91% of cases are sequenced, 36% of B.1.617.2 cases were associated with recent travel, yet the recent doubling in cases across all regions of Denmark suggests that B.1.617 and its sublineages has shifted to spreading beyond the household and close contacts of identified travellers. As of the 30th of May, B.1.617.2 makes up 80% of all recorded B.1.617 cases making up 0.5% of total cases (*27*). Germany and the Netherlands are sequencing a smaller proportion of cases (10% and 3% respectively) and report a peak in B.1.617 cases in early May. In Germany, B.1.617 cases account for 2% of cases in late May, with less than 0.5% in the Netherlands. This is consistent with travel-related importation and a delay in reporting of B.1.617.2. Our findings suggest that further growth of B.1.617.2 is likely in those countries, should patterns in England be repeated.

The UK vaccine uptake is very high: over 75% of the population have received one dose, and over 50% have received both doses at the time of writing (early June 2020). A key question for the UK’s continued easing of social distancing measures is the effectiveness of currently used vaccines against B.1.617.2. However, here we demonstrated that S-gene positive cases were initially in distinct subpopulations, both regionally and in terms of age groups. This observation could partly explain the difference in growth rates between S-gene positive and S-gene negative cases, if behavioural patterns imply differences in contact rates (*24, 25*) or propensity for larger gatherings (*26*) that is not dependent on the biology of B.1.617.2. However, the initial differences observed in age distributions have now converged to the background distribution, suggesting that the S-gene positive, and thus B.1.617.2, growth rate has stabilised to a higher value than B.1.1.7. The observation that age-related incidence is initially perturbed has the potential to affect vaccine effectiveness studies (*27*) if confounders are not completely accounted for. Current analysis of B.1.617.2 severity from Public Health England suggests that, when age is controlled for, the risk of hospitalisation remains increased in infections caused by B.1.617.2 (*23*). Our analysis highlights the complex and dynamic relationship between space, time, and age of cases in this outbreak, and the challenges in fully controlling for these variables when estimating phenotypic properties of emerging variants.

Despite remaining limitations (Supplementary Material Section 8), the United Kingdom has a -well-established disease surveillance programme and has the ability to undertake detailed epidemiological studies that establish phenotypic properties of globally circulating variants that are often not possible in locations where variants emerge or are first identified (*29*). Our work provides a deeper understanding of the effect of SARS-CoV-2 variant dynamics that need to be accounted for when estimating transmissibility, severity (*4–6*) and vaccine escape potential (*27*), and there is an ethical imperative to continue these efforts because the results have global policy implications.

## Supporting information

NHS REC ethics statement

## Data Availability

Data is available to researchers with data sharing agreements in place with Public Health England.

## Acknowledgments

We thank the members of JUNIPER consortium for helpful comments on the manuscript.

UKRI through the JUNIPER consortium (grant number MR/V038613/1), LDa LDy, JRG, MJK, CO, LG, FS, LP, DJP

MRC (grant number MC/PC/19067), LDa, RC, EBP, MJK

MRC (grant number MR/V009761/1) EMH, MJT, LDy, MJK

MRC (unit programme number MC UU 00002/11), DJP

EPSRC (EP/V051555/1), LDa, DW

EPSRC through the The Alan Turing Institute, grant EP/N510129/1), LDa, DW National Institute of Health Research UK: EBP,

Pfizer through investigator-led grants on respiratory tract infections. LDa

Wellcome Trust and the Royal Society (grant 202562/Z/16/Z), LP, HS and CO

Alexander von Humboldt Foundation, HS

## Author contributions

Conceptualization: RC, LDa, LDy, EBP, MJK

Methodology: RC, LDa, LDy, EBP, MJK, DW, CO, FS, LP,

Investigation: all

Visualization: RC,CO,LG,

Funding acquisition: LDa, MJK, LDy, EBP, JRG, MJT

Project administration: LDa, JRG, MJK, EBP

Supervision:LDa, JRG, MJK, EBP

Writing – original draft: LDa, RC, EBP, LDy, EMH, CO, HS

Writing – review & editing: All

## Supplementary Materials

### Materials and Methods

#### 1. Materials: Description of data sources

We estimated lineage-specific growth rates, age distributions and geographical spread using data provided by Public Health England (PHE) collected as part of the pandemic monitoring effort in England and provided to the Scientific Pandemic Influenza group on Modelling (SPI-M).

We use information on variants of concern (VOCs) or variants under investigation (VUIs) in the UK from four data streams: the positive SARS-CoV-2 cases line list; the S-gene line list detailing TaqPath test results; the VAM line list detailing variants of concern identified by genomic sequencing from COG-UK; and the CTAS (Contact Tracing Advisory Service) line list, detailing genomic sequencing results that are not variants of concern. Descriptive analyses of the frequency of variants of concern among patients who have a S-gene positive TaqPath test result were performed using all four data sources to create a combined genomic and S-gene data set, de-duplicated into unique episodes of infection. For the analysis of growth rates, and associated age distributions, we used only the positive cases line list and the S-gene line list, de-duplicated using 4 different methods, as detailed below, and excluding the last 4 days of case counts, which are subject to reporting delays, and may bias recent estimates of growth rates. In our main results we present findings based on unique episodes of infection and provide further sensitivity analyses in this section.

Here, we describe each dataset in turn and the data fields that are applicable to the sequencing and/or determining S-gene status of specimens. We later describe the pipeline to combine the multiple data sources.

##### Dataset 1: Variant line list (VAM)

This contains a list of specimens sequenced by the Covid Genomics UK Consortium (COG-UK), provided by Public Health England (PHE), that were genomically confirmed as VOCs or VUIs; as of 20 May in the UK there were 5 VOCs and 7 VUIs (Table 1 of PHE SARS-CoV-2 variants of concern and variants under investigation in England: Technical Briefing 10; (*1*)). Additionally, each record included the traveller status of the individual (Traveller, Contact of Traveller, Not travel-associated, Refused or Uncontactable, Awaiting information). The VAM is deduplicated to one VOC/VUI call per person. If multiple VOCs per person are identified (rare), non-B.1.1.7 (e.g., B.1.617.2) is prioritised over B.1.1.7 which is prioritised over unclassified.

##### Dataset 2: CTAS line list

This contains genomic information collected through the Contact Tracing Advisory Service (CTAS) and included traveller status. It lists all sequences that could be linked to the contact tracing system, including sequences that were not VOCs or VUIs. CTAS line list was the only source to contain sequences that were neither VOCs or VUIs.

##### Dataset 3: “Pillar 1 and Pillar 2 line lists”

These are lists detailing the first case for an individual (i.e. they are deduplicated) within Pillars 1 and 2 of the UK SARS-CoV-2 mass testing programme. Pillar 1 encompasses virus testing in PHE laboratories and NHS hospitals for those with a clinical need, and health and care workers. Pillar 2 contains records of virus tests for the wider population. It records the ethnicity, age and location (to coarse-grained geographic level) for each positive case. In addition, each line list record has a categorical value for “Asymptomatic_indicator” which details the symptomatic status of the individual on the date of testing for community (Pillar 2) tests. A value of “N” indicates the person declared symptoms at the time of testing; and “Y” can be interpreted as tests conducted for screening purposes (i.e. asymptomatic testing).

##### Dataset 4: “S-gene line list”

For Pillar 2 tests performed using the ThermoFisher TaqPath system, these supplied RT-PCR cycle threshold (Ct) values and their classification according to whether the S-gene target of the TaqPath assay failed to amplify. The TaqPath assay is a multiplex test designed to target three distinct regions of the SARS-CoV-2 genome (N, ORF1ab, S). Each record in the S-gene line list was classified as either S-gene negative, S-gene positive or equivocal, according to these Ct criteria: S-gene negative - N<=30 Ct; S undetected; ORF<=30 Ct; (also referred to as S-gene dropout) and S-gene positive - N<=30 Ct; S<=30 Ct; ORF<=30 Ct; (also known as “triple-positive”), and equivocal - other combinations. Tests taken during recovery are frequently equivocal when CT values rise.

##### 1.1 Processing I: case numbers for growth rate estimates

The S-gene line list is provided on a per-test basis, and some people have multiple S-gene test results. This can be the result of repeated testing during a single episode of infection or multiple episodes of repeated infection, and there is some ambiguity in this. We ensured each infection episode was counted no more than once in the following four ways:

###### 1) Selecting only cases for whom the S-gene test result is within 4 days of their first ever positive result for that patient (“first infection”)

This is simple but potentially biases the data, under-representing people who have had multiple infections or for whom the first test in an infection episode was done in a lab that does not use TaqPath tests and followed up with a TaqPath test later in the infection.

###### 2) Selecting only cases with a first positive TaqPath test result for an individual (“first taqpath”)

By taking the first ever TaqPath test result for an individual we maximise the number of individuals we identify but potentially bias the data because the date of the sample may not be completely representative of the date of onset of disease, if the patient was initially diagnosed by a laboratory that does not use TaqPath, or by a lateral flow device. Also, cases with repeated infections, separated by a long time, would be excluded from analysis.

###### 3) Selecting only the last positive TaqPath result for an individual (“last taqpath”)

By taking the last ever TaqPath test result we also potentially bias the sample by making the time point of infection more recent, in the subset of patients who have had multiple TaqPath test results. However, if these test results are widely separated in time, this strategy may be appropriate as it will pick up the most recent infection episodes.

###### 4) Inferring continuous infection episodes based on delays between tests (“infection episodes”)

This strategy involves grouping all known test results, both TaqPath and non TaqPath, together into continuous episodes, containing sequences of positive test results separated by 28 days or fewer (or 56 days in the event of an equivocal S-gene test result). Each episode may contain multiple test results, and if there are TaqPath results within the episode that are positive, and there are no negative S-gene results, the episode is deemed to be caused by a S-gene positive infection. Conversely if there are negative TaqPath test results within an episode and no positive results the episode is deemed to be caused by a S-gene negative infection. It is categorised as equivocal if TaqPath test results are all equivocal, and unknown if there are no TaqPath test results for that episode.

This strategy has the benefit that we correctly identify the onset date of the episode regardless of when the TaqPath testing was done in an episode, and potential episodes of re-infection are detected. Compared to other methods it may appropriately result in earlier infection dates if they have had a few tests done in laboratories that do not use TaqPath tests.

##### 1.2 Processing II: S-gene and genomic sequencing case data

Assembling a linked data set for genomic variant and TaqPath result informs our analysis of case numbers. The data processing pipeline has five main steps:

###### Step 1: Create a joint confirmed sequence case list from the CTAS and VAM linelists

- From the VAM line list we took proven VOC and VUI cases (i.e. confirmed through sequencing).
- From the CTAS line list we took proven non-VOC/VUI cases.
- We used the following groups: B.1.351, P.1 & P.2, B.1.617.1, B.1.617.2, other VOC/VUI.
- We included the B.1.1.7 and B.1.525 variants within the “other VOC/VUI” category.

###### Step 2: Gather unsequenced S-gene positive cases from the S-gene line list

- We added entries in the sequenced case list from Step 1 with a FINALID not present to the group of unsequenced S-gene positive cases from the S-gene positive line list, deduplicated using the infection episodes strategy.

###### Step 3: Construct a combined list of sequenced VOCs and VUIs (from Step 1) and unsequenced S-gene positive cases (from Step 2)

- We include both confirmed and suspected cases with VOCs/VUIs of interest.

###### Step 4: Using Pillar 1 and Pillar 2 line lists, determine if a case was asymptomatic undertaking a test as part of a screening process

- Check for match of FINALID and specimen_date fields in the Pillar 1 and Pillar 2 line lists and the Step 3 combined list.
- Extract value from the Asymptomatic_indicator field: U for Pillar 1; N for Pillar 2 and symptomatic; Y for Pillar 2 and test conducted for screening purposes (i.e. asymptomatic testing)
- Non-matches could occur if it was not the first sample that was sent for sequencing and we could not match the date. This may happen either when a case is a reinfection or if multiple specimens were taken. We recorded these instances as “Unknown” for asymptomatic indicator.

###### Step 5: Using Pillar 1 and Pillar 2 line lists, determine ethnicity, age and location of a case

- Check for a match of the FINALID field between the Pillar 1 and Pillar 2 line lists and the Step 3 combined list.
- If a match is found, extracted data from the fields for ethnicity, age and patient location (LTLA level).
- We assume these remain unchanged throughout the study period, acknowledging this will not account for people aging or moving residence.

#### 2. Methods: Instantaneous growth rate estimation

We measured the growth rate of the S-gene positive and S-gene negative cases to assess the potential of VOCs. The growth rate describes the exponential rate at which cases are growing or declining in a given area. As opposed to the widely used reproduction number (*2*), the growth rate can be estimated directly from data and provides a direct measure of the speed of growth of cases regardless of whether they derive from direct transmission or other sources such as importation. It is therefore a more reliable measure to investigate trends when prevalence is low and importation may be significant. From a given growth rate, classical methods allow to compute the corresponding reproduction number using estimates of the generation time distribution (*3–5*), but this implicitly makes the assumption that the observed growth is driven entirely by local transmission. Additionally, the generation time distribution of a new variant is often hard to infer from the scarce available data and may in general be different from that of previously circulating variants.

Especially in situations of low prevalence, as at the time of writing (June 2021), outbreaks can be very heterogeneous across the country. For this reason, in addition to looking at aggregated national figures, which could average out areas seeing rapid spread with others still in decline, we estimated the growth rates in different smaller-scale geographies independently. These local outbreaks can be indicative of the speed of growth of a national epidemic were a variant to become widespread across the country, although conditions at the local scale may not translate to larger geographies. Further, estimating reliable trends in the growth rate when looking at small scale geographies is challenging due to the very low number of cases involved and that the instantaneous growth rate is undefined when no cases are observed in a given time period. To mitigate this problem and handle the uncertainty with the data, we applied four independent methods to estimate the instantaneous growth rate, each of them with different assumptions, as described below.

##### Generalised Additive Model method

To estimate growth rates, we adapt a generalised additive model (GAM) where the number of cases on day, *t*,^*I*(*t*)^, is assumed to be given by *I*(*t*) ∝ *exp*(*s*(*t*)) for some smoother function *s*(*t*). We use a log link and a penalised spline as implemented in the R package mgcv (*6*). The over-dispersed noise inherent in both disease dynamics and surveillance data motivates the use of a negative binomial error structure, and a day-of-the-week fixed effect is added to capture daily variability within a 1-week period. The number of knots used by the spline is fixed as one twentieth the length of the time-series (for time-series shorter than 200 days the default number of knots is used) to avoid over smoothing the data or losing signal in the noise. The instantaneous local growth rate is then the time derivative of the smoother. The GAM can lead to boundary effects from the choice of smoother, so the most recent central estimates may not be reliable. The growth rate is assumed independent for each geographical area and case definition considered, where case definitions include S-gene positive and S-gene negative. The model used is an extension of the model developed by Pellis et al. 2021 (*7*).

##### Gaussian Process method

The growth rate is estimated independently for each geographical area. For a given area i, the method assumes that the daily count of cases is distributed as a negative binomial function with risk parameter *μ_it_*, where *t* is a day index. We decompose the log-relative-risk parameter into a Gaussian process (GP) and a weekday random effect: log (*μ_it_*) = *g_t_* + *w_t_*, where wt has a log-gamma distribution with shape 0 and rate 0.01, and gt is a GP with a Matérn covariance function with v=3/2, length scale l and precision tau. The hyperparameter l is assigned a log-normal prior with mean 1 and precision 1, while tau has a log-normal distribution with mean −3.5 and precision 100. The growth rate is calculated as the first derivative of the GP. To remove fine-scale fluctuations, the derivative is approximated using a centred difference approximation. The model is implemented using the R-package INLA, where the GP is obtained as the weak solution of a stochastic partial differential equation (*8, 9*).

##### Poisson regression method

The growth rate is estimated for every day using a generalised linear model, where the number of cases on day, *t*^*I*(*t*)^, is assumed to be given by *I*(*t*) ∝ *exp* (*rt*) which fits a quasipoisson model to the surrounding 8 weeks of case counts, as a direct estimate of as the growth rate. When considering dates within the most recent 4 weeks there is less data to estimate the growth rate on, so the most recent estimates are both less reliable and more representative of the growth rate in the past. As quite a large window of data is used, this method is slow to respond to step changes in the instantaneous growth rate.

##### Poisson Bayesian method

The same approach as the poisson regression was also implemented in Bayesian framework using an observation level random effect, rather than quasipoisson error distribution to account for the overdispersion. This was fitted in the R package brms v 2.15.0 (*10*). The priors used were a normal (mean 0, standard deviation 5) prior on the intercept of the model, a standard normal prior on the growth rate (***r***), and an exponential (lambda=1) prior on the standard deviation of the observation random effect distribution. As in the poisson regression method, the surrounding 8 weeks of case counts were used, and the caveats about the most recent 4 weeks of data apply. This method was used to validate the main estimates in Figure 2 of the paper and is not being used in the sensitivity analysis.

Figures S2.1 and S2.2 show estimates of the instantaneous growth rate obtained using the three different methods. For sensitivity, we tested different datasets obtained with the four alternative deduplication processes (1–4) presented in Section 1.1. Consistent results across different methods and data cleaning processes give us confidence on the general trends of S-gene positive and negative cases.

**Figure S2.1.**
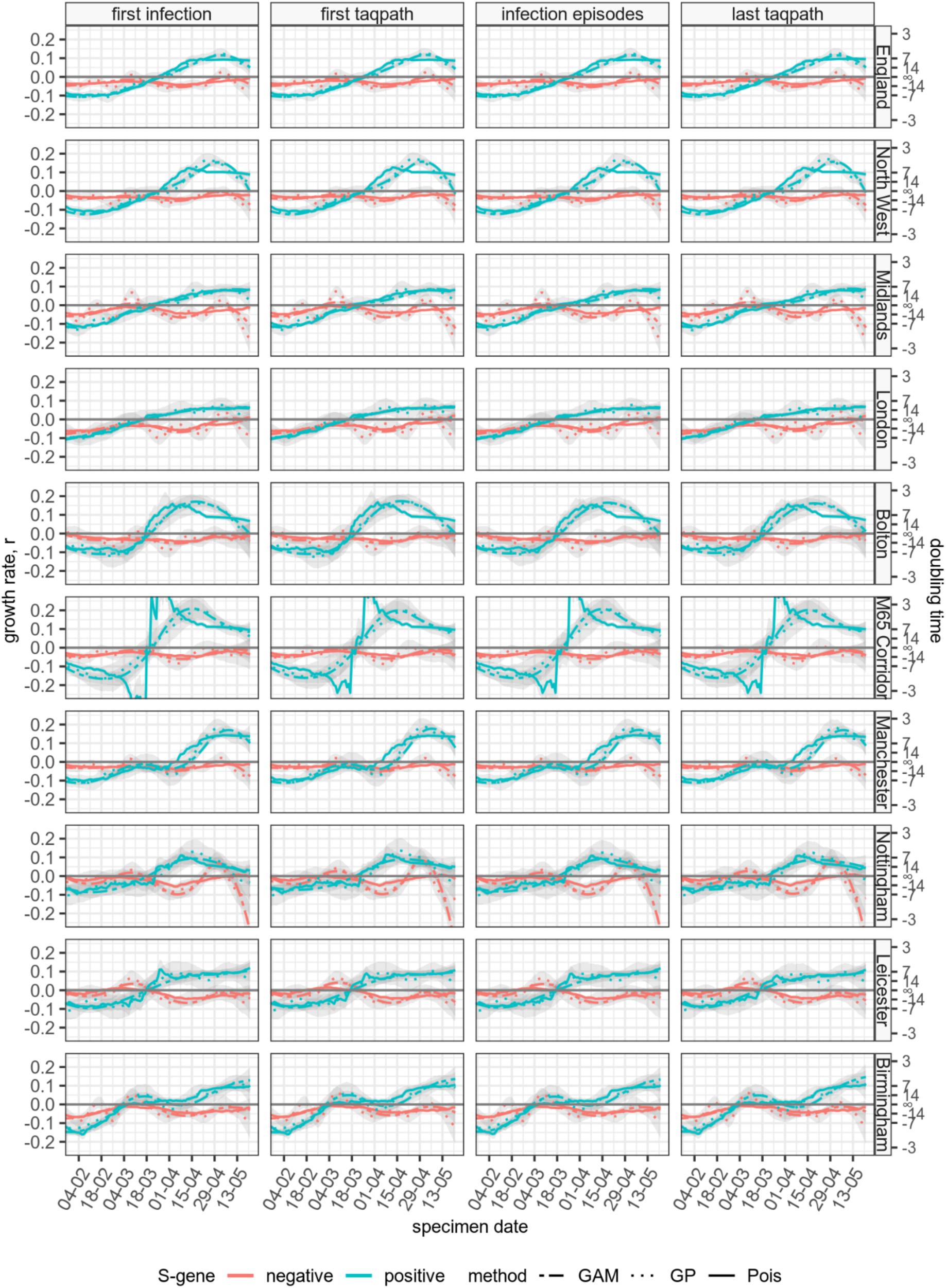
Instantaneous growth rates and doubling times for all regions estimated using three of the different methods explained in Section SI2 (General additive model, Gaussian Process and Poisson regression) and data obtained using the four different deduplication strategies explained in Section 1.2.

**Figure S2.2.**
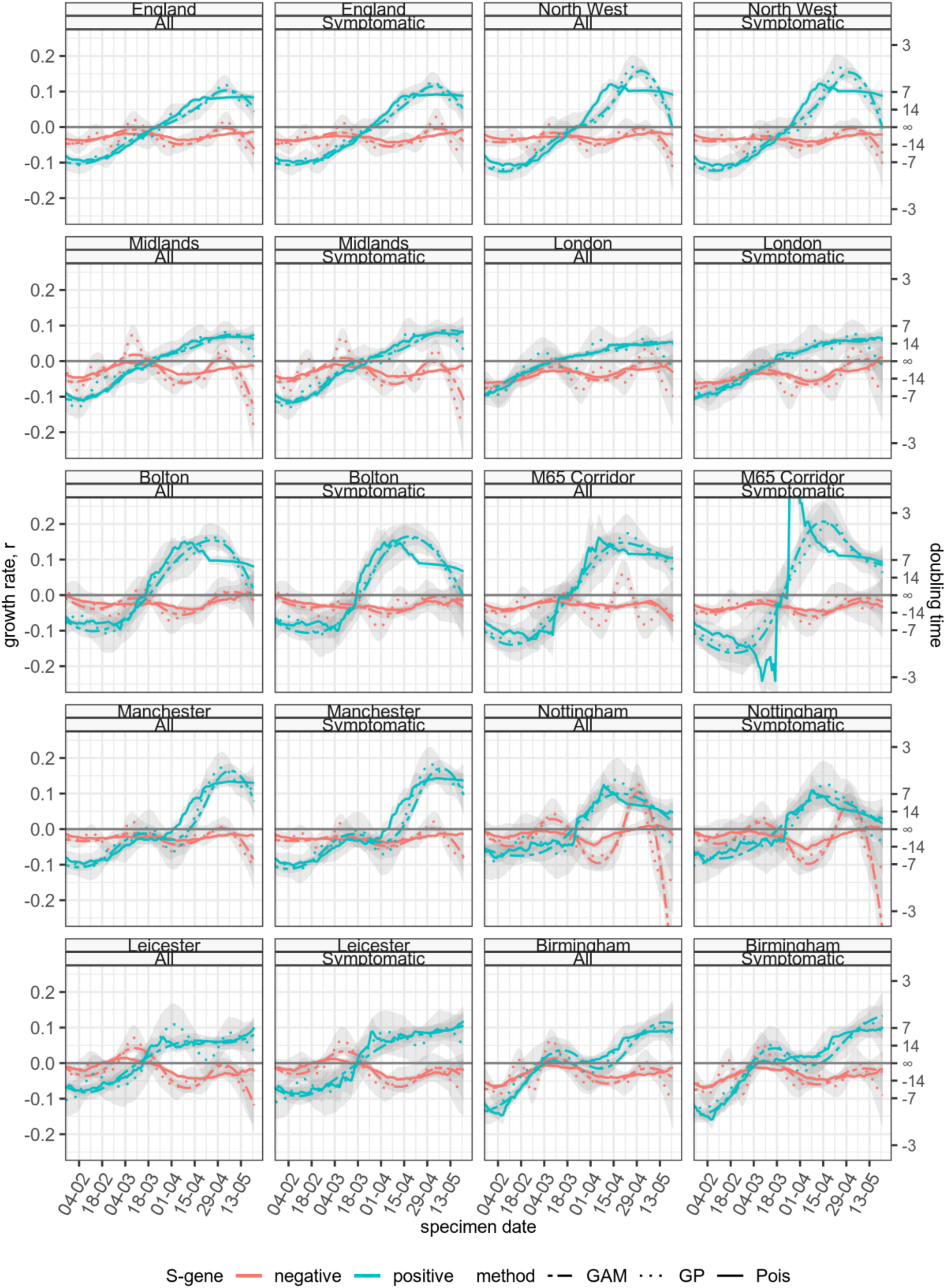
Instantaneous growth rates and doubling times for all regions estimated using three of the different methods explained in Section 2 (General additive model, Gaussian Process and Poisson regression) using all cases, versus the subset of cases described as “symptomatic”.

#### 3. Methods: Comparison of the distribution of cases by age

Individuals’ transmission patterns typically depend on age, and interactions between different age groups can drive epidemics. Using empirically-derived age mixing matrices describing those interactions, next generation matrices can be calculated (*11*), which map the age distribution of infected individuals in a population to the age distribution after one generation of infections. As the epidemic progresses, the age distribution of infected individuals converges to the dominant eigenvector of the next generation matrix (*12*), provided interaction mixing and transmission patterns remain constant, and assuming the same generation time distribution for all ages. Under the assumption that mixing and transmission patterns (i.e. relative susceptibility and infectivity by age) are similar for all variants of a pathogen up to a multiplicative constant describing an overall increase or decrease in transmissibility, we would expect cases across different variants to have the same age distribution, regardless of whether the epidemic is growing or declining. However, when a new variant emerges, it may emerge preferentially within and between certain age groups, though as the variant-level epidemic progresses the age distribution should eventually converge to the same dominant eigenvector. In the context of B.1.617.2, many cases arrived in the UK through travel, possibly resulting in an age bias in B.1.617.2 cases aligned with the age distribution of travellers. Following these seeding events, we may observe three primary outcomes: continued growth through importation of cases, continued growth through community transmission, or local extinction of the variant. If we observe continued growth through importation, the age distribution of cases will reflect those of the imported cases. If continued growth occurs through community transmission, the age distribution of cases should gradually shift towards the dominant eigenvector, which describes the mixing and transmission patterns in the community. If local extinction occurs, the cases should die out, and we again expect a distribution indistinguishable from the dominant eigenvector, but without a corresponding growth in cases. Therefore, by studying the age distribution of different variants in tandem with the growth rate, we can gain insight into whether there is community transmission.

To investigate the age distribution of cases, we use the Wasserstein (or Kantorovich) distance metric (*13, 14*). Given a metric space M provided with a metric d, the 1st Wasserstein distance, or ‘earth mover’s distance’, or Mallow’s distance, intuitively describes the minimum cost of transforming one probability distribution to another. Let X and Y be random variables with distributions P and Q in R^d^, respectively. The Wasserstein distance between P and Q, ^*M_p_* (*P, Q*^), can be defined as the minimum of the expected difference between X and Y, taken over all joint probability distributions F for (X,Y) such that the marginal distribution of X is P and the marginal distribution of Y is Q, i.e.,

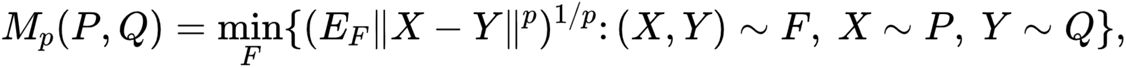

We consider the 1st Wasserstein distance, defined for p=1. Some of the advantages of the Wasserstein distance rather than other measures, e.g., the Kullback–Leibler divergence, in the context of age distributions, are: 1) the Wasserstein distance satisfies the properties of a metric (e.g., it is symmetric); 2) it is well defined for probability distributions with different support; 3) it is appropriate for information like age where the distance between different values has meaning.

To ensure conclusions are not driven by the choice of distance metric, we also use the Kolmogorov-Smirnov distance measure. This is defined as the largest absolute difference between two cumulative distribution functions, i.e.,

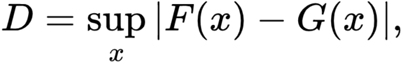

where F and G denote the cumulative distribution functions of the probability distributions under study. The Kolmogorov-Smirnov test is conceptually simple but suffers from some drawbacks including that it focuses on the point of maximal difference rather than measuring more global differences between distributions. We here use this measure only as an additional validation of the tests performed using the Wasserstein distance.

To compare two empirical distributions (e.g., the observed age distributions of S-gene positive versus S-gene negative cases), we consider the null hypothesis that the two samples are drawn from the same age distribution. We do this by using a permutation test: we combine the two samples into a single distribution, from which we randomly draw 1000 permutations that split this single distribution into two samples, with the respective sample sizes of the original two samples. For each pair of permutation samples, we calculate their (Wasserstein or Kolmogorov-Smirnov) distance. These distances represent possible values that could be obtained if the two samples were drawn from the same distribution. From the 1000 permutation distances, we can calculate percentiles in order to obtain a confidence region. We use the latter to determine whether the observed samples are likely to have been drawn from the same distribution: if the distance calculated from the original samples lies within the confidence region, then there is insufficient evidence to reject the null hypothesis that the two samples were drawn from the same distribution, at the corresponding confidence level.

**Figure SI 3.1:**
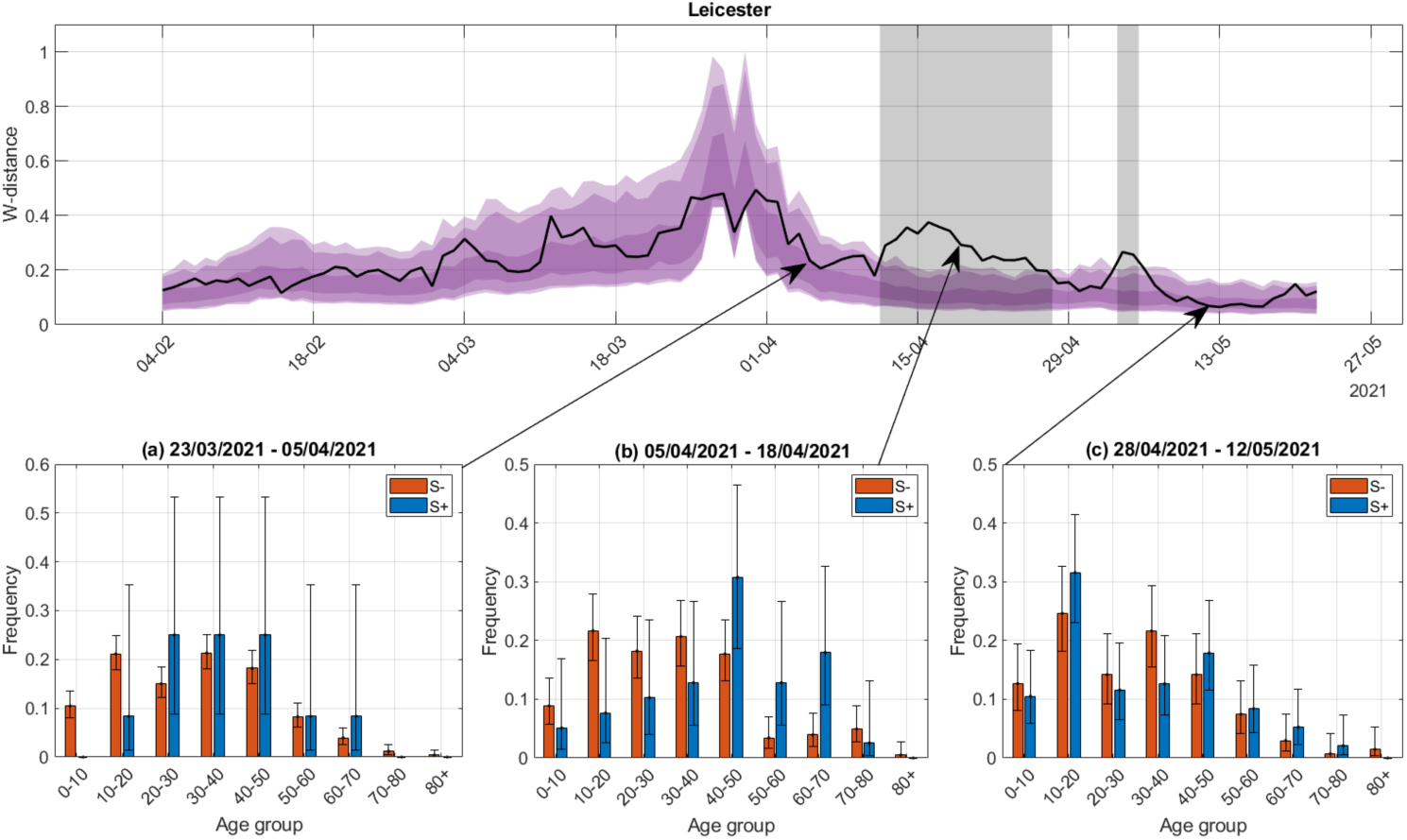
Comparing the Wasserstein time-series to snapshots of the observed age distributions for S-gene positive and S-gene negative cases in Leicester. This figure illustrates how the Wasserstein distance relates to the observed distributions.

Figure S3.1 focuses on Leicester as an example. The bottom three panels show a snapshot of the age distributions at given points in time. Panel (a) shows the age distribution of cases between 23/03/2021 and 05/04/2021. Visually comparing the S-gene positive and S-gene negative age distributions, there does not seem to be a notable difference between the two, at least within the binomially distributed uncertainty bounds. This is in agreement with the fact that the Wasserstein distance for the corresponding date range falls well within the confidence region. The second panel shows the age distributions between 05/04/2021 and 18/04/2021. There is now a more evident difference in the observed age distributions, with S-gene positive cases occurring in older individuals. The corresponding Wasserstein distance is outside the confidence region, confirming the significant difference between the two age distributions. The final panel shows the distributions for cases between 28/04/2021 and 12/05/2021. Visually the two age distributions appear similar again, and the Wasserstein distance is back within the confidence region, so there is no evidence to suggest a significant difference between the distributions. Therefore, the relative position of the computed Wasserstein distance and the confidence region obtained by a permutation test allows quantification of the visual relationship between the age distributions and indicates whether such distance is significant based on the sample sizes involved. This is particularly important when either one or both sample sizes are low and visual inspection may be difficult. In this case, the Wasserstein distance can be very high even when the two samples are drawn from the same distribution, leading to large confidence regions.

To investigate temporal changes in the relationship between the two age distributions, we considered the age distributions over a rolling time window. We opted for a 14-day time window, as this ensured that there were sufficient cases to get some insight into the age distribution, whilst still having a fine-grained temporal resolution. Using a longer window would result in a temporal correlation of long duration, so short-term, but substantial perturbations to the age distributions could be missed. Performing this for the regions of interest, for the Wasserstein distance, gives the results shown in Figure 3 in the main paper. To verify that our conclusions regarding comparisons between the data streams are not driven by the assumptions of the Wasserstein distance, in Figure S3.2, we show the same analysis for the Kolmogorov-Smirnov distance alongside the Wasserstein distance. Comparing the results, both metrics suggest similar conclusions. Therefore, our conclusions are not driven by the choice of model.

**Figure S3.2:**
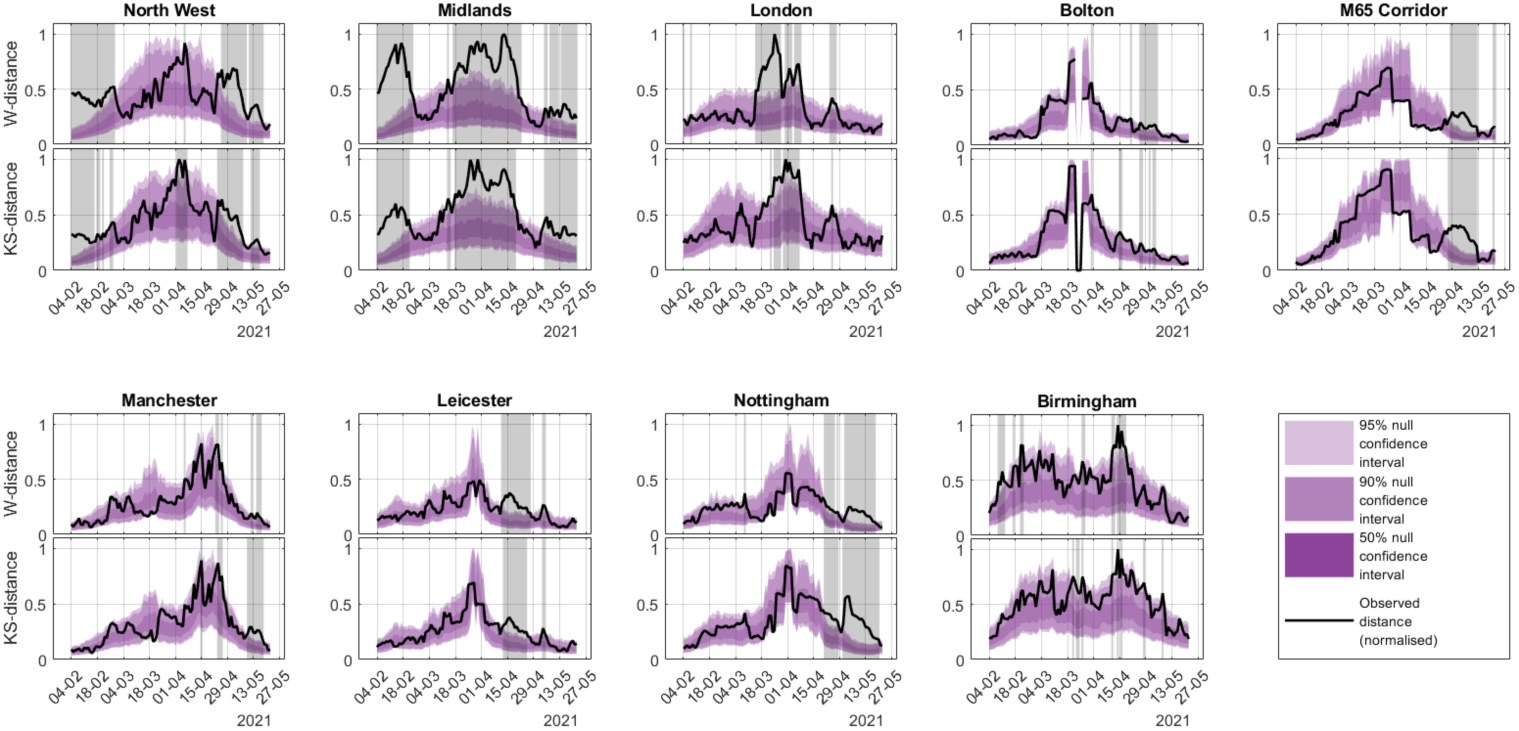
Comparison of the Kolmogorov-Smirnov and Wasserstein distances between the S+ and S- age distributions, in the areas of concern. This considers the age distribution among a two-week rolling aggregation of cases, plotted by last swab date in the two-week window. The black curve indicates the distance between the two age distributions. The purple shaded region is generated through 1000 Monte Carlo samples of a permutation test, and indicates the confidence region for distance metrics if the two samples were drawn from the same distribution. The grey shaded regions indicate dates where the distance metric falls outside of the confidence region, denoting a significant difference between the age distribution of S-gene positive and S-gene negative cases.

For comparison, we show the age distributions in the regions of interest below:

**Figure S3.3:**
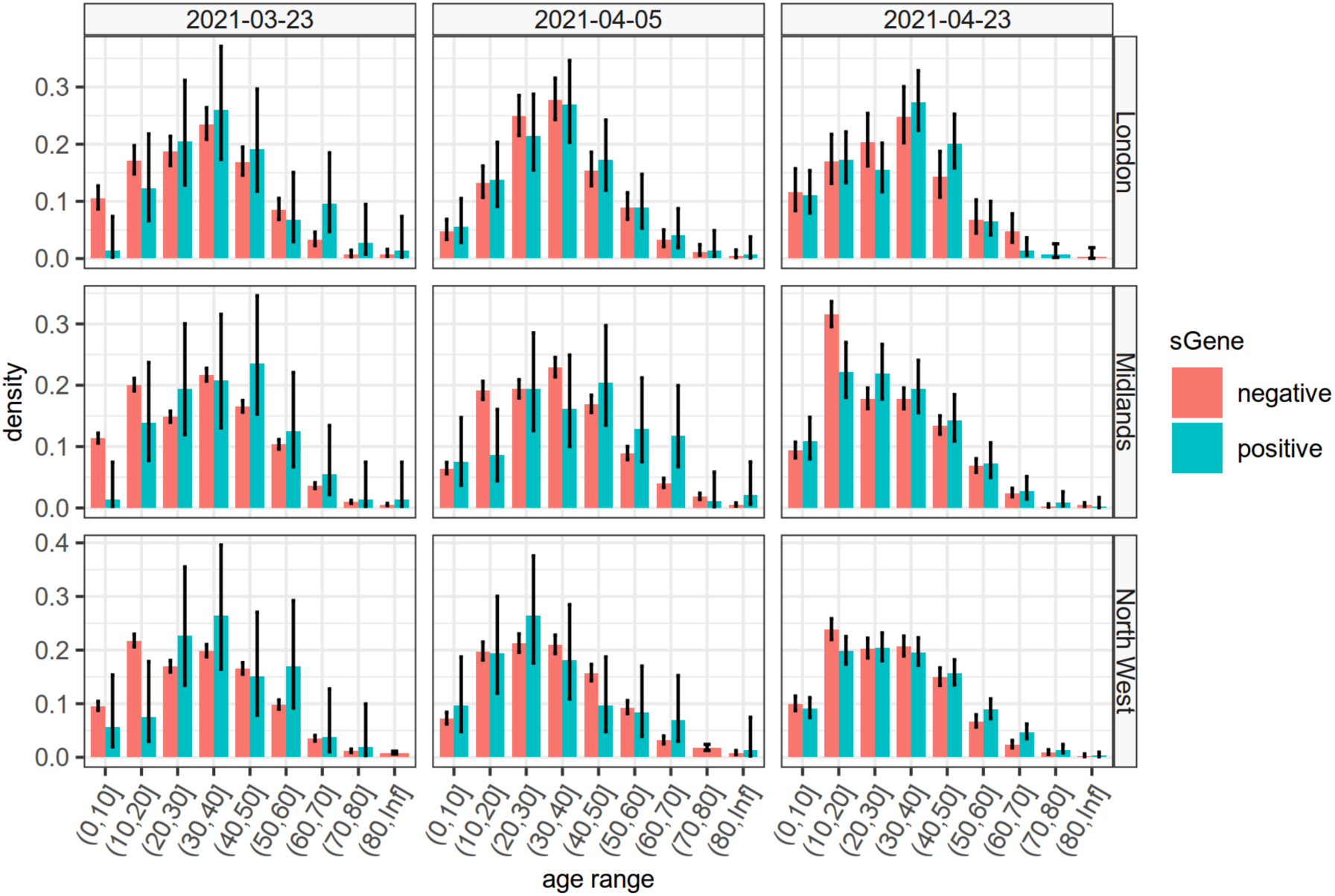
Age distributions of S-gene positive and negative detected COVID-19 cases over a two week period from the dates shown in column headers, for 3 NHS regions of England with good TaqPath test coverage.

**Figure S3.4:**
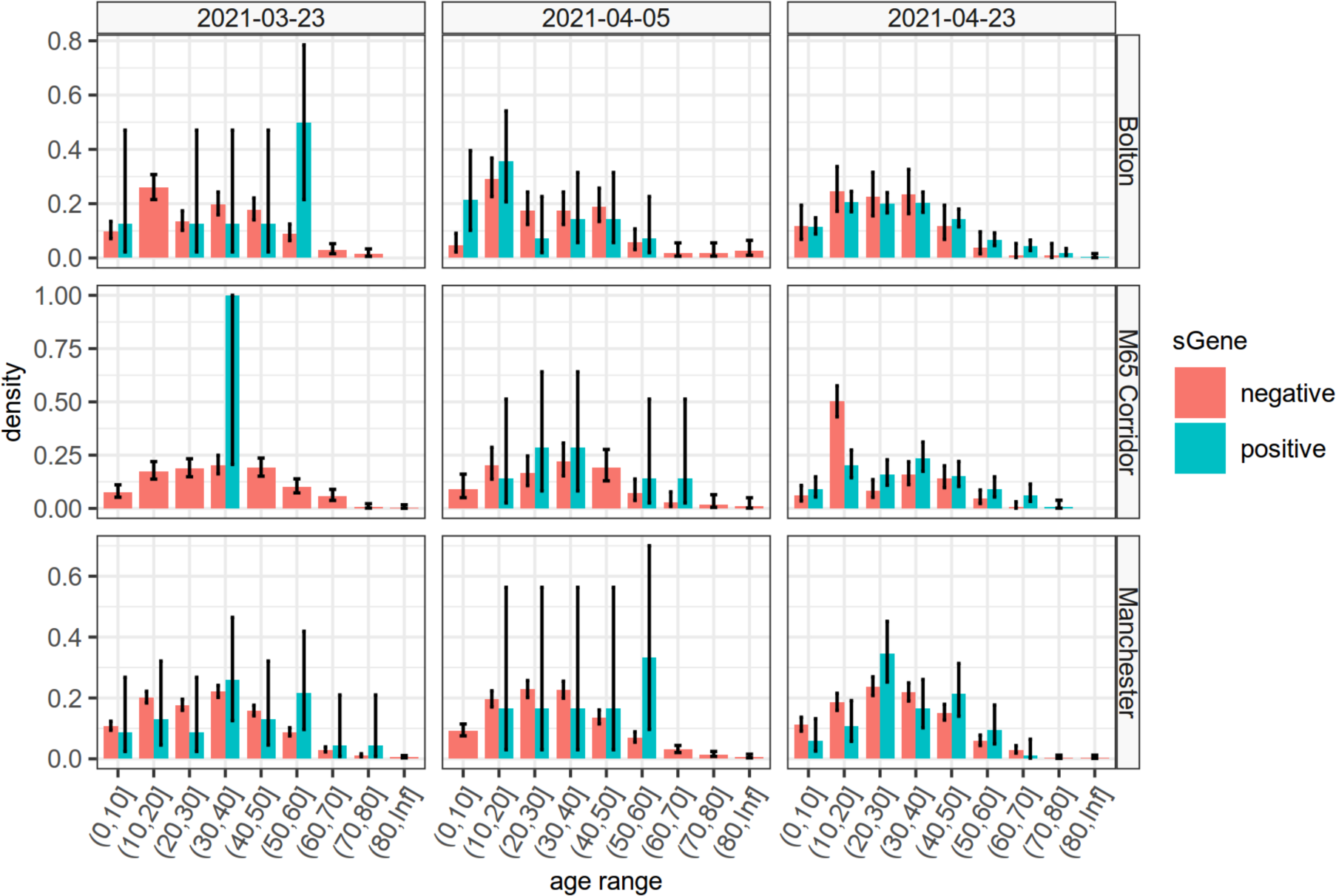
Age distributions of S-gene positive and negative detected COVID-19 cases over a two week period from the dates shown in column headers, for 3 smaller geographic regions of the North West of England, associated with S-gene positive COVID-19 outbreaks.

**Figure S3.5:**
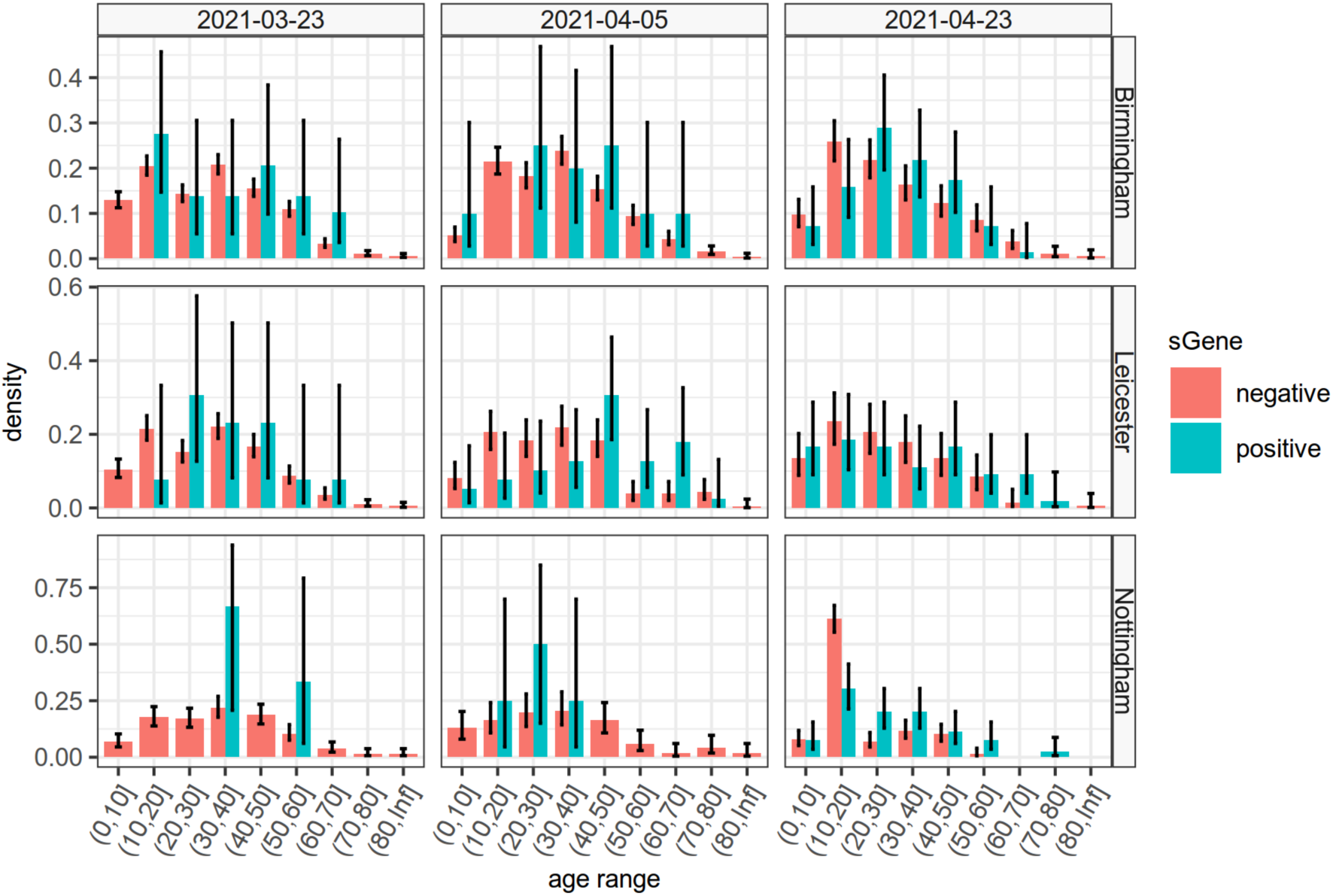
Age distributions of S-gene positive and negative detected COVID-19 cases over a two week period from the dates shown in column headers, for 3 smaller geographic regions of the Midlands of England, associated with S-gene positive COVID-19 outbreaks.

##### Age distribution of cases among travellers

Cases in travellers may not reflect the age distribution of community acquired cases. To investigate this, we compare the subset of PCR positive cases in confirmed travellers to all PCR positive cases (Figure S3.6). Both samples consider cases between 28/02/2021 and 24/05/2021, to ensure the time frames are comparable. The age distribution of traveller cases is skewed towards older individuals, suggesting that the perturbations seen in the age distribution of S gene positive cases are likely to be caused by imported cases.

**Figure S3.6:**
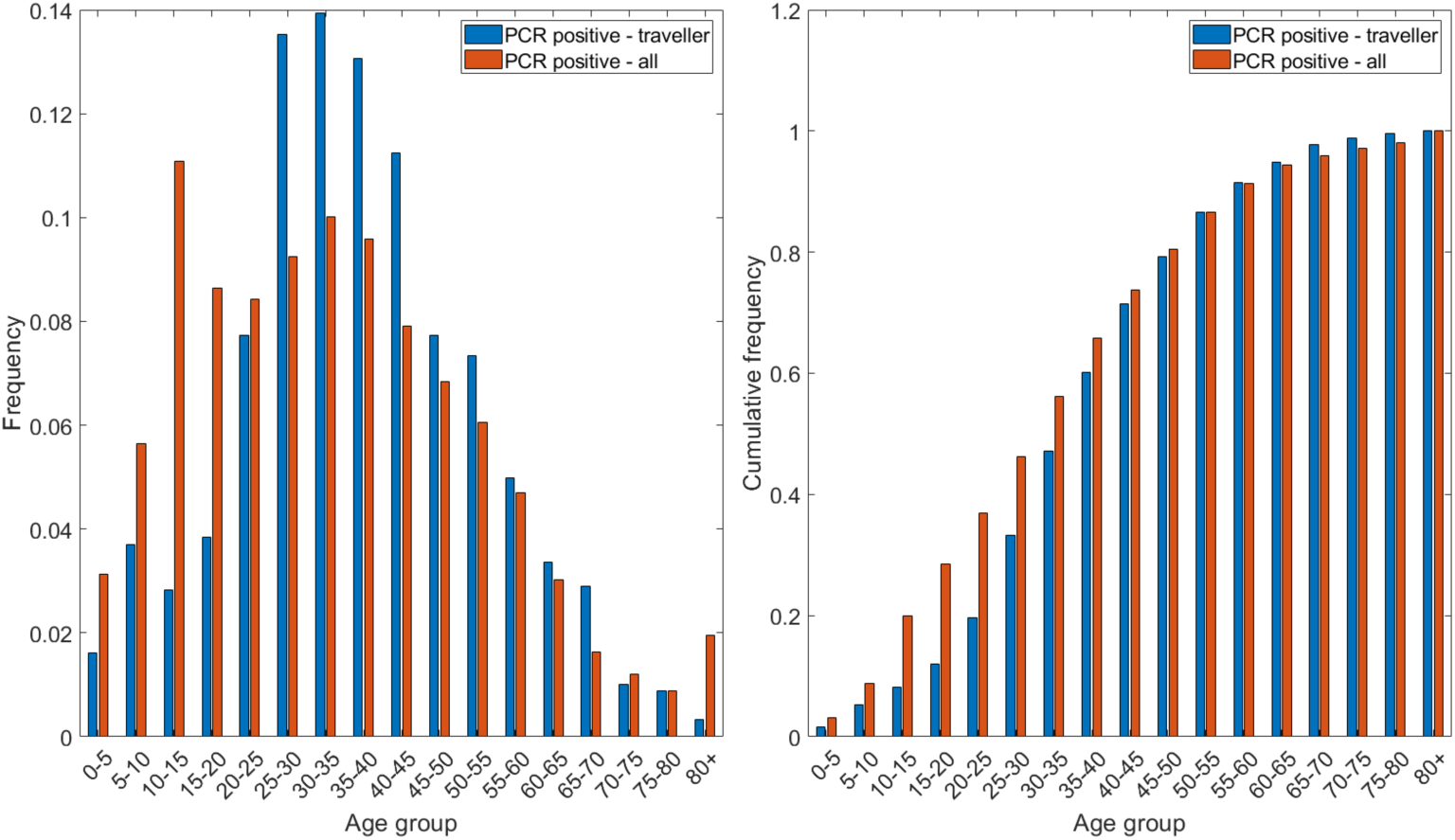
Age distribution of cases in travellers versus all cases. The left plot shows the proportion of cases in each age group, and the right plot shows the cumulative frequency up to each age group. From the left plot, the traveller age distribution appears skewed towards older individuals. The right plot confirms this, with the age distribution of all cases having a substantially higher cumulative frequency for lower ages.

#### 4. Regions analysed

Here we detail the definition of the regions that were used as units of analysis. In Table S4.1, the area of analysis, the name and the corresponding Office of National Statistics codes are listed. The regions were chosen to represent various different geographical levels where we have relatively good coverage both in terms of TaqPath S-gene status and also in terms of sequencing results. At the top level we analysed the whole of England, at the next level we analysed the NHS regions of the North West, the Midlands, and London. The North West and Midlands have the best overall TaqPath coverage in England, whereas London has a lower level of coverage and is included for comparison, (see Supplementary Material Section 6). The most granular regions were selected by focussing on areas where we see significant B.1.617.2 outbreaks, in the areas with good TaqPath coverage. These areas including Bolton, the M65 Corridor, and Manchester in the North West, and Nottingham, Leicester and Birmingham in the Midlands, were areas identified where numbers of sequenced B.1.617.2 were high, from the analysis presented in Section 5. In identifying these low level regions, we ensured that the areas were areas where the vast majority of sequenced S-gene positive cases were B.1.617.2, and where the B.1.617.2 cases were most clearly associated with community transmission, by investigating regions where the majority of cases were identified as symptomatic (rather than screening), where there was no clear evidence of high levels of case importation, and where the ethnicity and age distributions of cases were heterogeneous (see Section 7).

**Figure S4.1.**
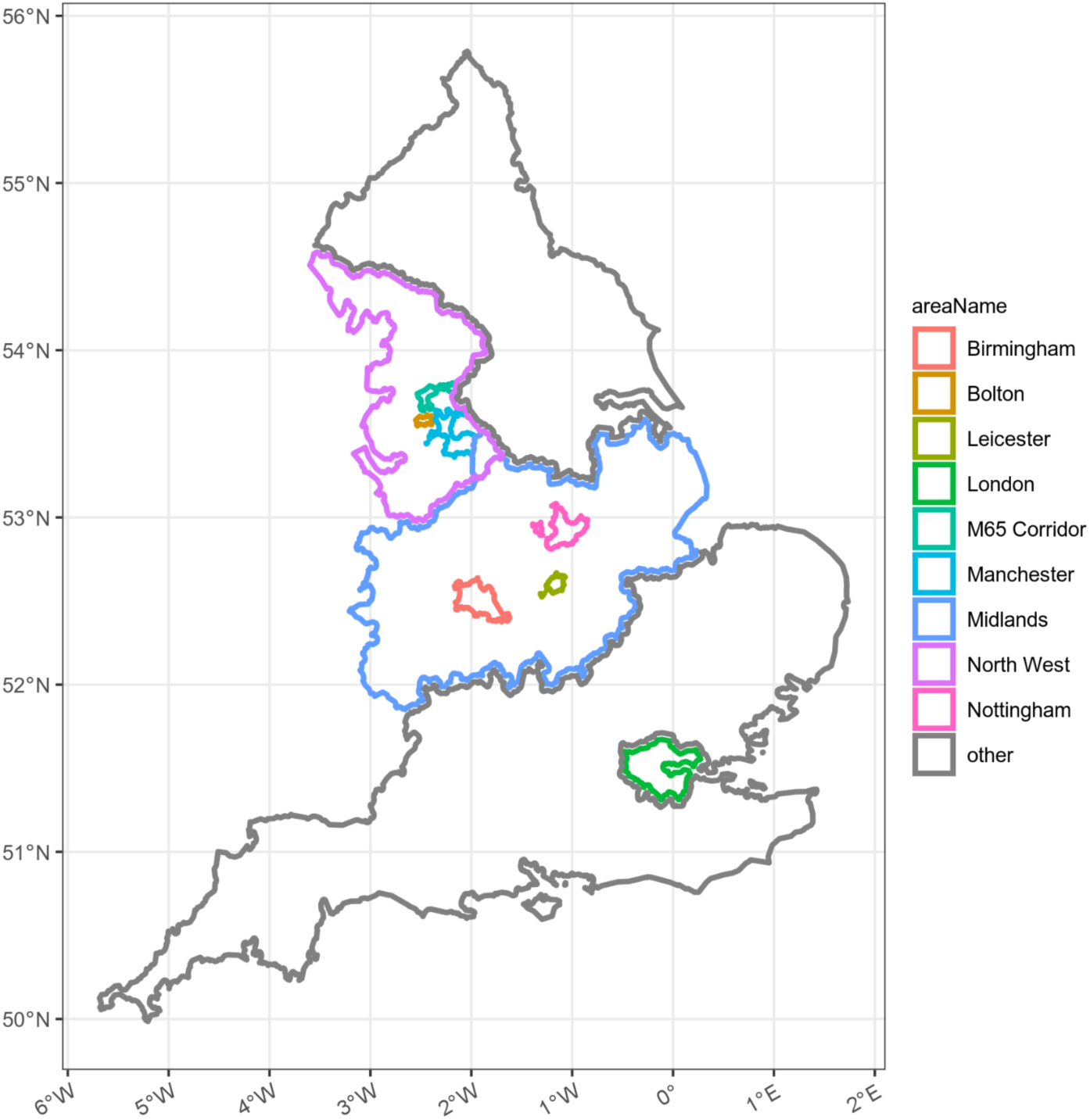
Mid and lower level regions analysed and presented in the main paper.

**Supplemental Table S4.1.** UK Administrative Lower Tier Local Authority regions associated with regions analysed.

| Area of analysis | LTLA code | Name |
| --- | --- | --- |
| Bolton | E08000001 | Bolton |
| M65 Corridor | E06000008 | Blackburn With Darwen |
|  | E07000120 | Hyndburn |
|  | E07000117 | Burnley |
| Nottingham | E06000018 | Nottingham |
|  | E07000036 | Erewash |
|  | E07000173 | Gedling |
|  | E07000176 | Rushcliffe |
| Leicester | E06000016 | Leicester |
|  | E07000129 | Blaby |
|  | E07000135 | Oadby and Wigston |
| Birmingham | E08000025 | Birmingham |
|  | E08000031 | Wolverhampton |
|  | E08000028 | Sandwell |
|  | E08000027 | Dudley |
|  | E08000029 | Solihull |
|  | E08000030 | Walsall |
| Manchester | E08000003 | Manchester |
|  | E08000006 | Salford |
|  | E08000008 | Tameside |
|  | E08000007 | Stockport |
|  | E08000005 | Rochdale |
|  | E08000002 | Bury |
|  | E07000037 | High Peak |

#### 5. Geographical distribution of S-gene positive cases, sequenced B.1.617.2 infections and B.1.1.7 sequences

**Figure S5.1:**
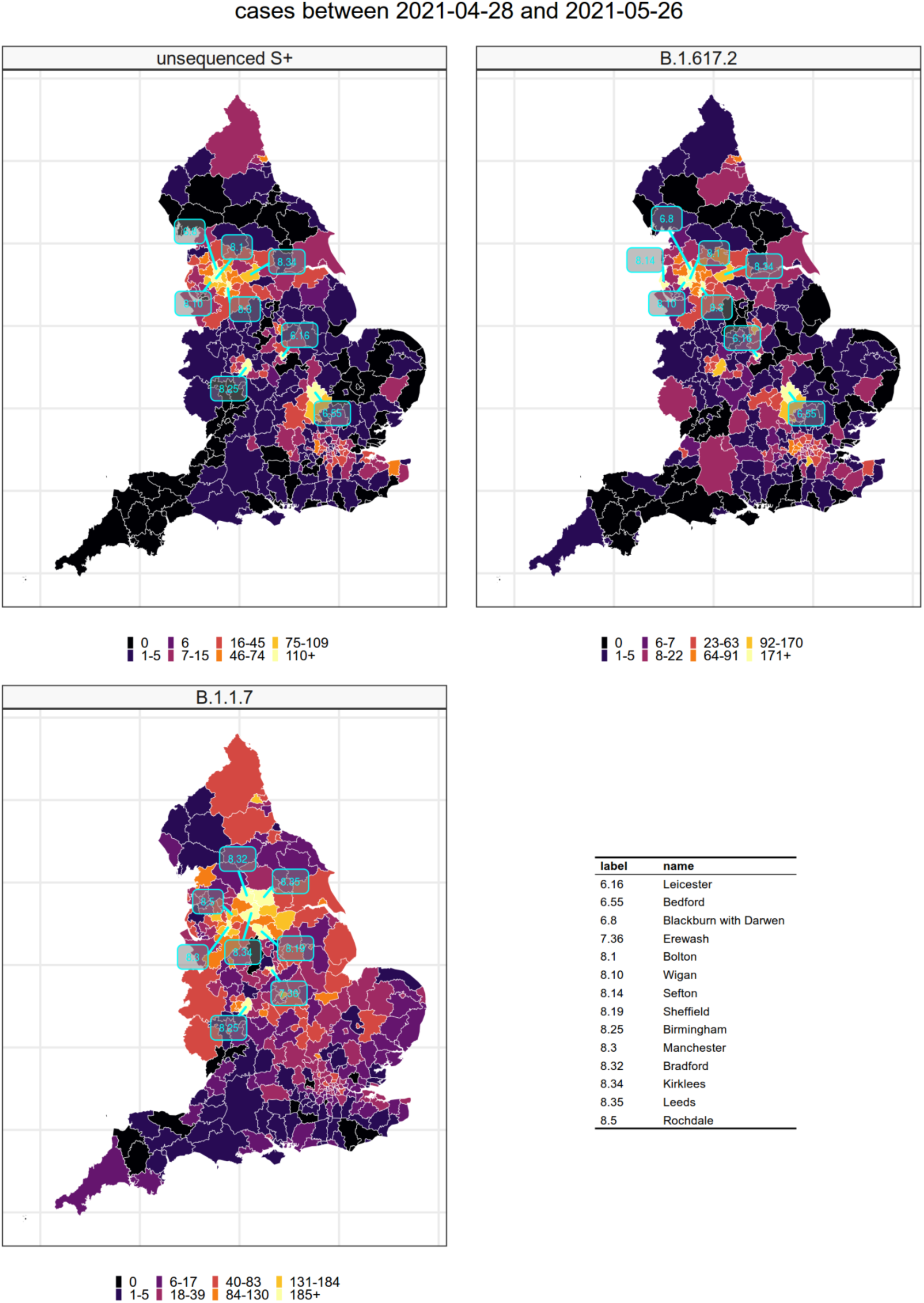
Geographical co-location of unsequenced S-gene positive cases and confirmed B.1.617.2 cases, in comparison to confirmed B.1.1.7 cases in the 4 weeks ending on the 26th May 2021, in England as a whole.

**Figure S5.2:**
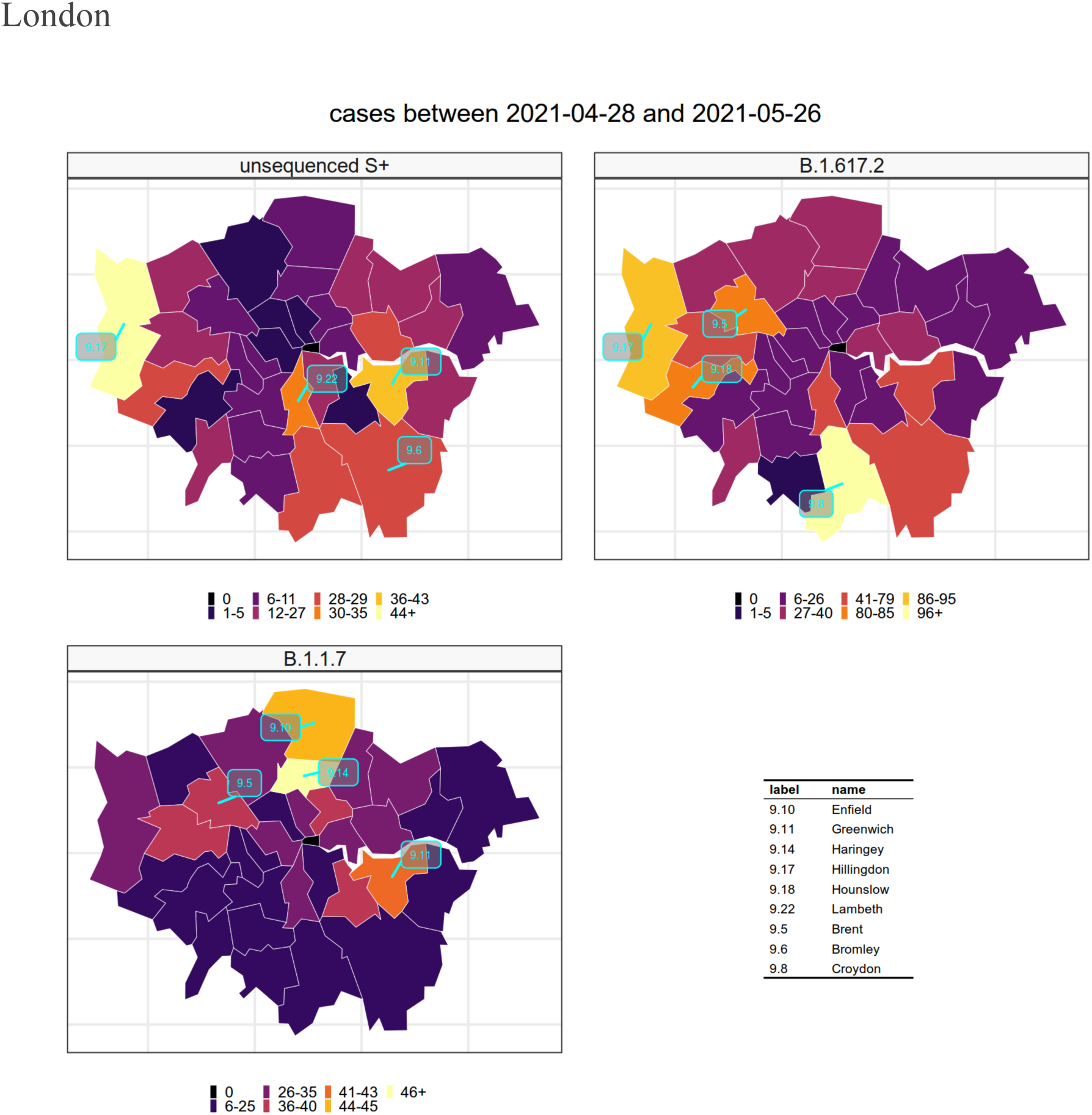
Geographical co-location of unsequenced S-gene positive cases and confirmed B.1.617.2 cases, in comparison to confirmed B.1.1.7 cases in the 4 weeks ending on the 26th May 2021, in the NHS region of London.

**Figure S5.3:**
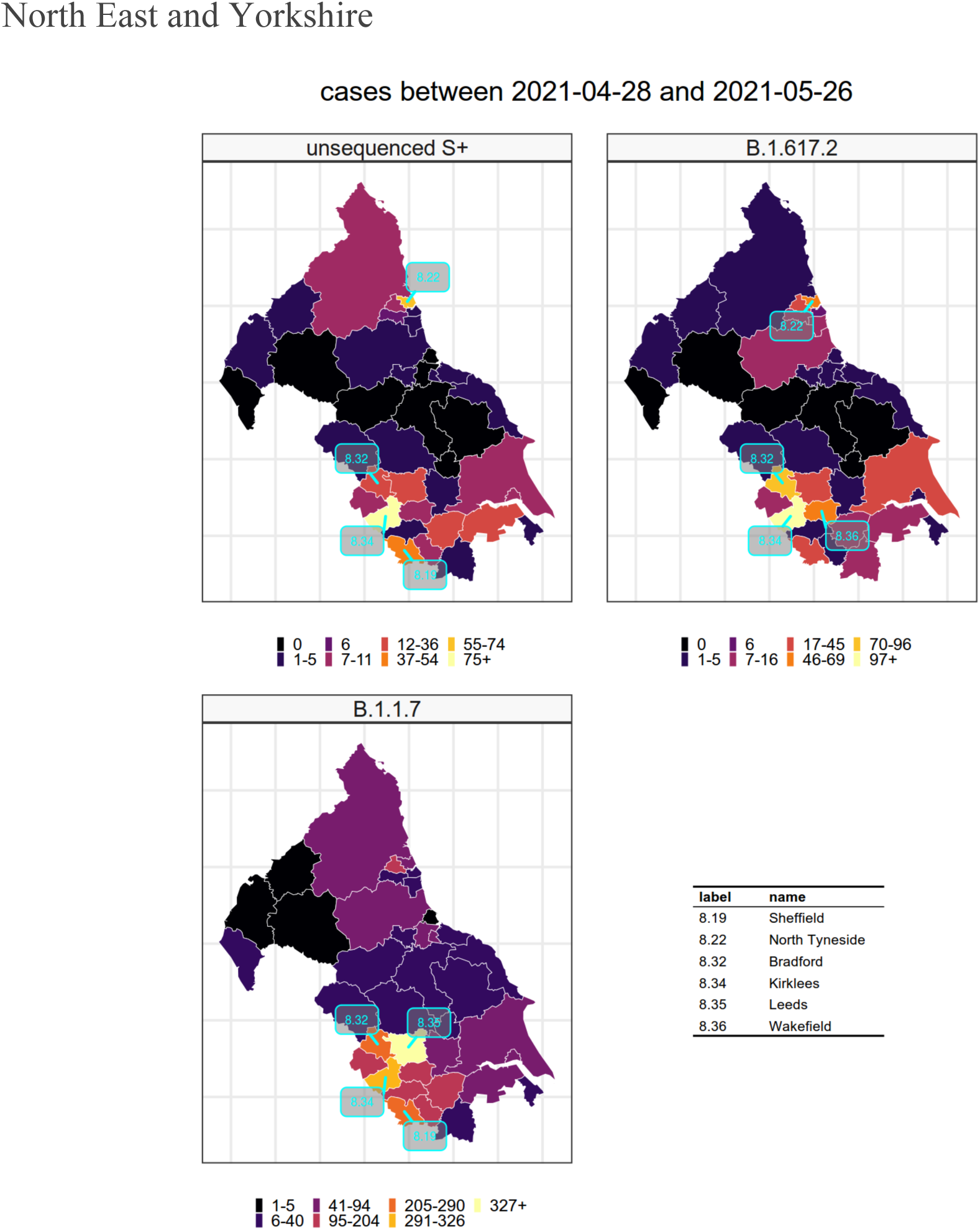
Geographical co-location of unsequenced S-gene positive cases and confirmed B.1.617.2 cases, in comparison to confirmed B.1.1.7 cases in the 4 weeks ending on the 26th May 2021, in the NHS region of the North East and Yorkshire.

**Figure S5.4:**
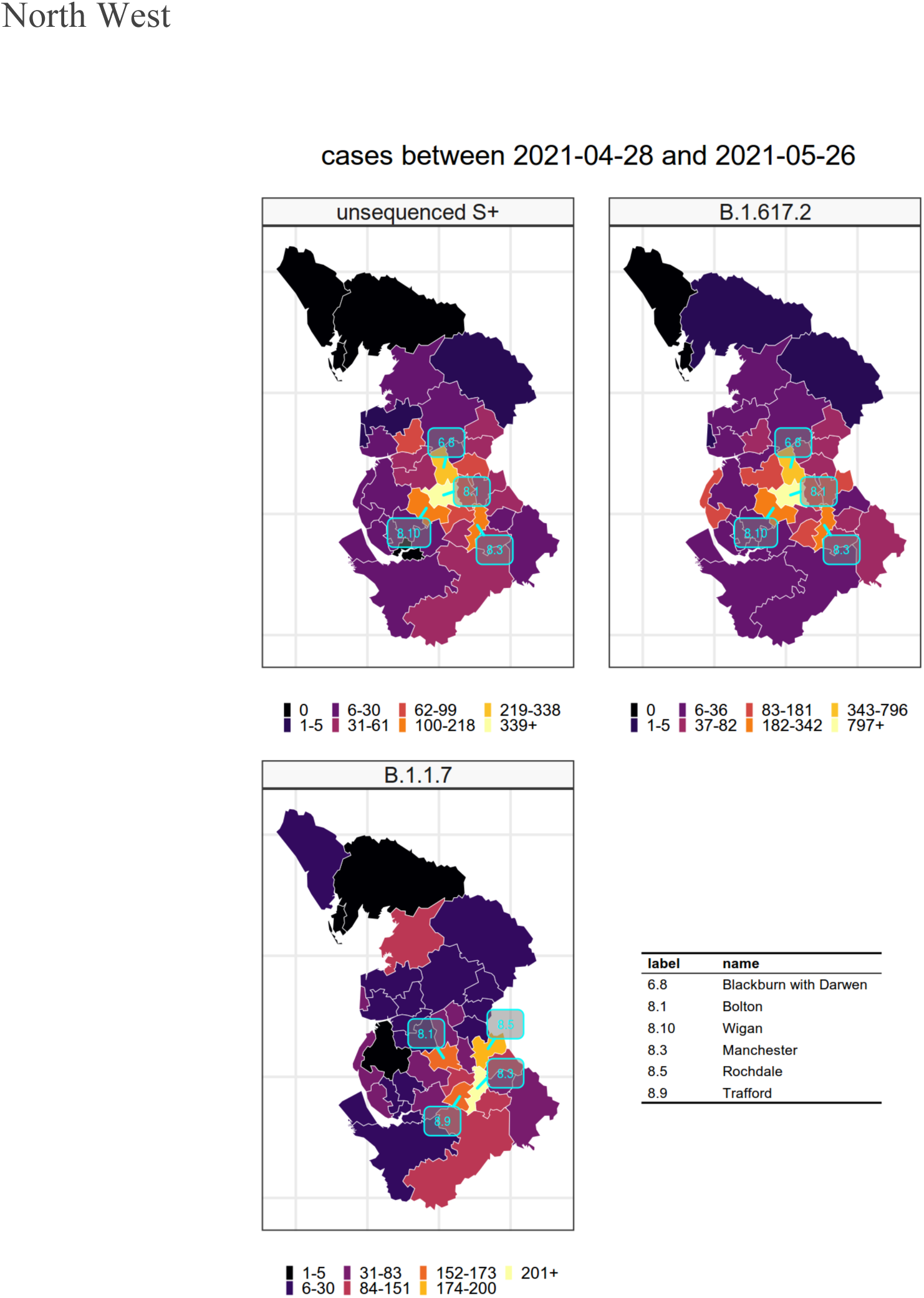
Geographical co-location of unsequenced S-gene positive cases and confirmed B.1.617.2 cases, in comparison to confirmed B.1.1.7 cases in the 4 weeks ending on the 26th May 2021, in the NHS region of the North West.

**Figure S5.5:**
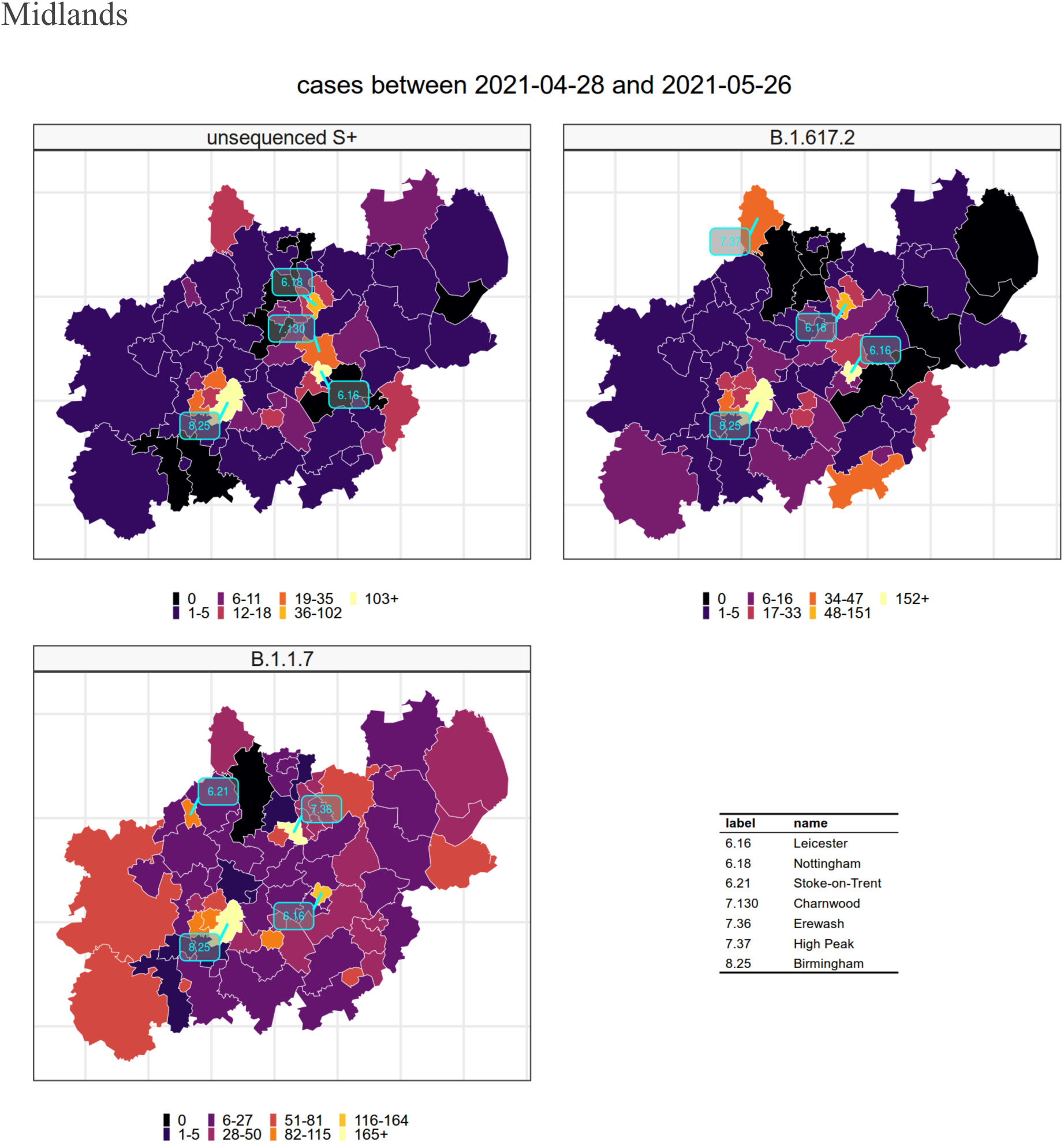
Geographical co-location of unsequenced S-gene positive cases and confirmed B.1.617.2 cases, in comparison to confirmed B.1.1.7 cases in the 4 weeks ending on the 26th May 2021, in the NHS region of the Midlands.

**Figure S5.6:**
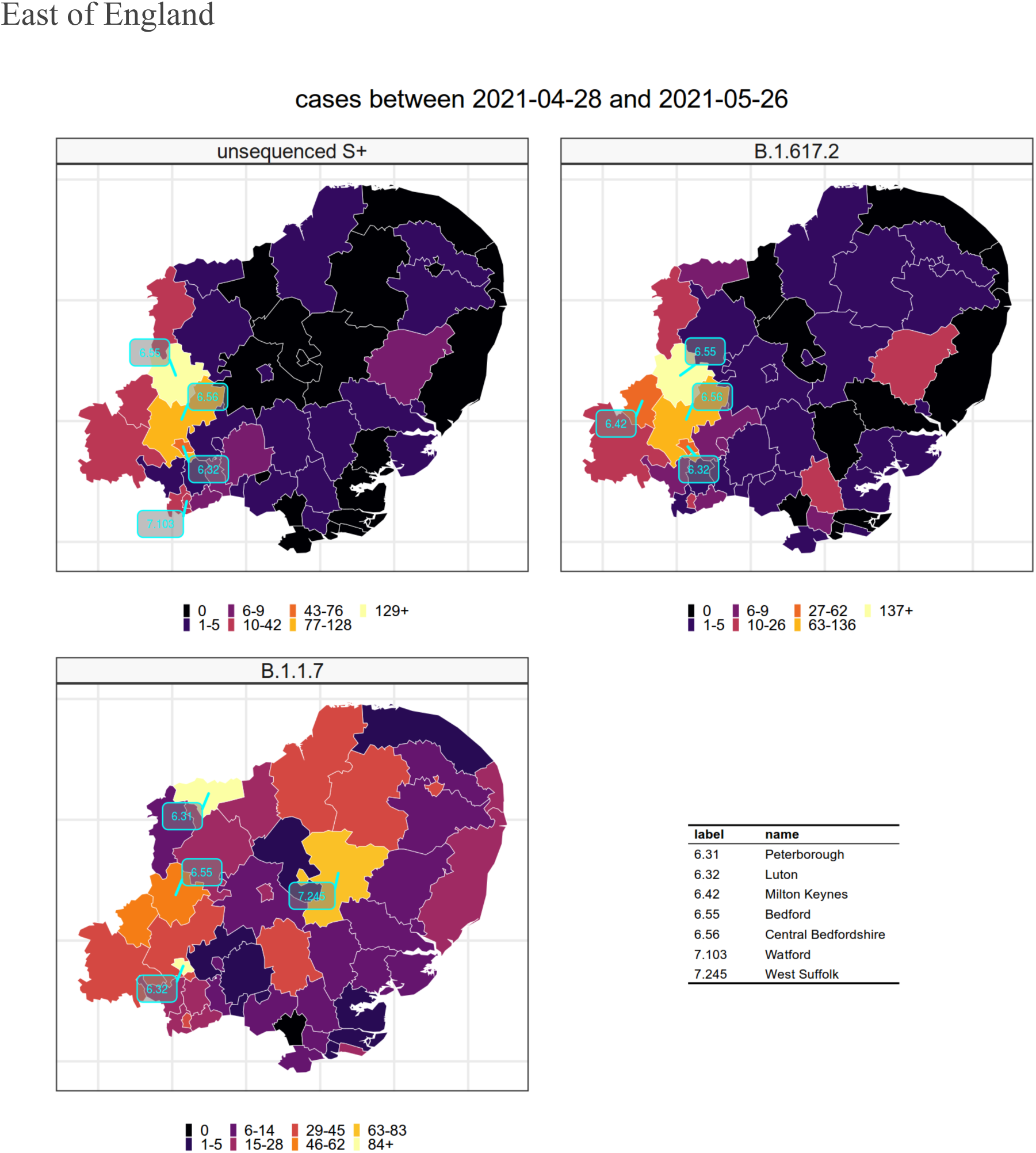
Geographical co-location of unsequenced S-gene positive cases and confirmed B.1.617.2 cases, in comparison to confirmed B.1.1.7 cases in the 4 weeks ending on the 26th May 2021, in the NHS region of the East of England.

**Figure S5.7:**
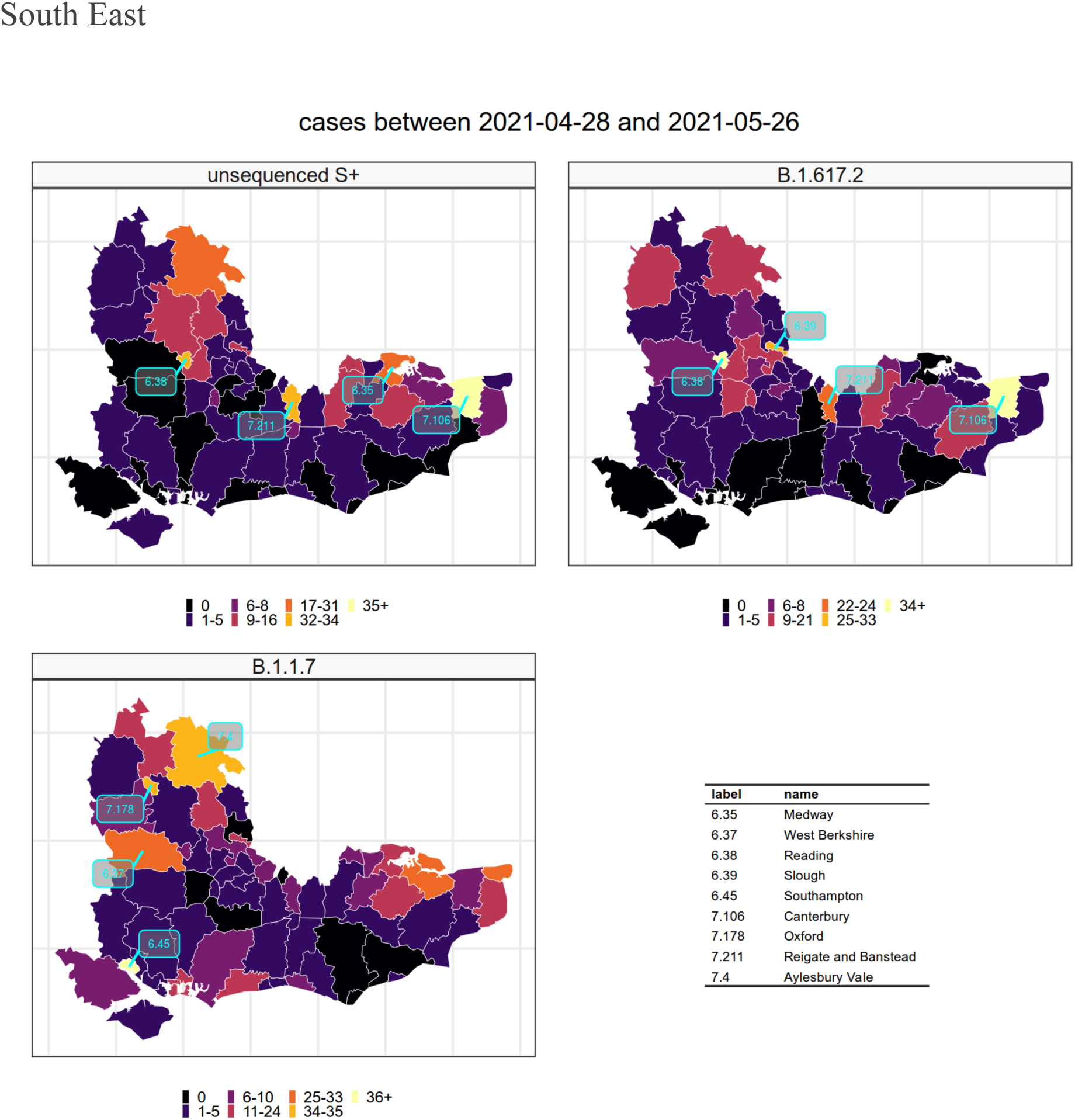
Geographical co-location of unsequenced S-gene positive cases and confirmed B.1.617.2 cases, in comparison to confirmed B.1.1.7 cases in the 4 weeks ending on the 26th May 2021, in the NHS region of the South East.

**Figure S5.8:**
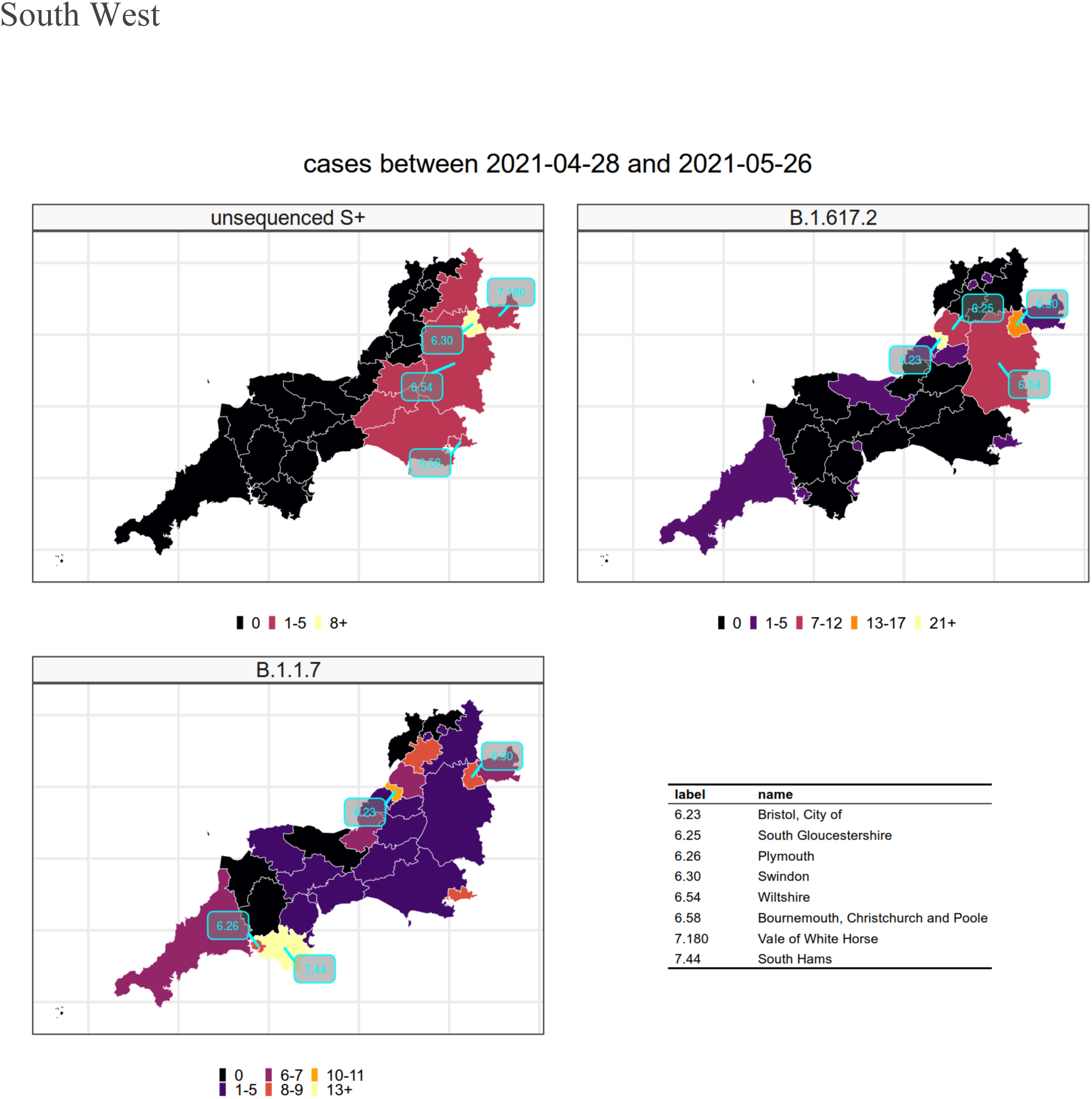
Geographical co-location of unsequenced S-gene positive cases and confirmed B.1.617.2 cases, in comparison to confirmed B.1.1.7 cases in the 4 weeks ending on the 26th May 2021, in the NHS region of the South West.

#### 6. S-gene TaqPath test coverage

The degree of testing using the TaqPath assay varies from lab to lab. Since 1st March 2021 coverage of the S-gene test has been more extensive in the regions which we identify as problematic. This may be the result of an acquisition bias.

Since 1st March 2021 the number of Pillar 2 positive cases varies substantially from region to region reflecting areas where the epidemic has taken more time to die down.

**Figure S6.1.**
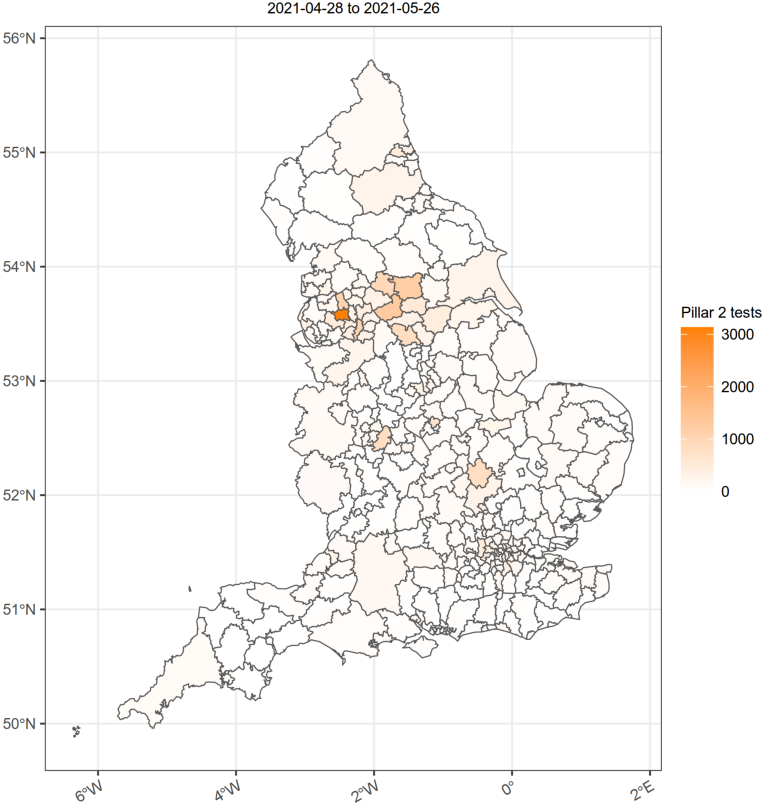
Pillar 2 positive cases by LTLA between 2021-04-17 and 2021-05-15.

The proportion of tests that are performed using the TaqPath testing system and therefore for which we will have S-gene results generally covers those areas which have had a lot of Pillar 2 testing. However, regions with low case numbers also tend to have low TaqPath coverage and hence the S-gene signal in these areas is unreliable. Overall the TaqPath coverage in London for example is only about 30%.

**Figure S6.2.**
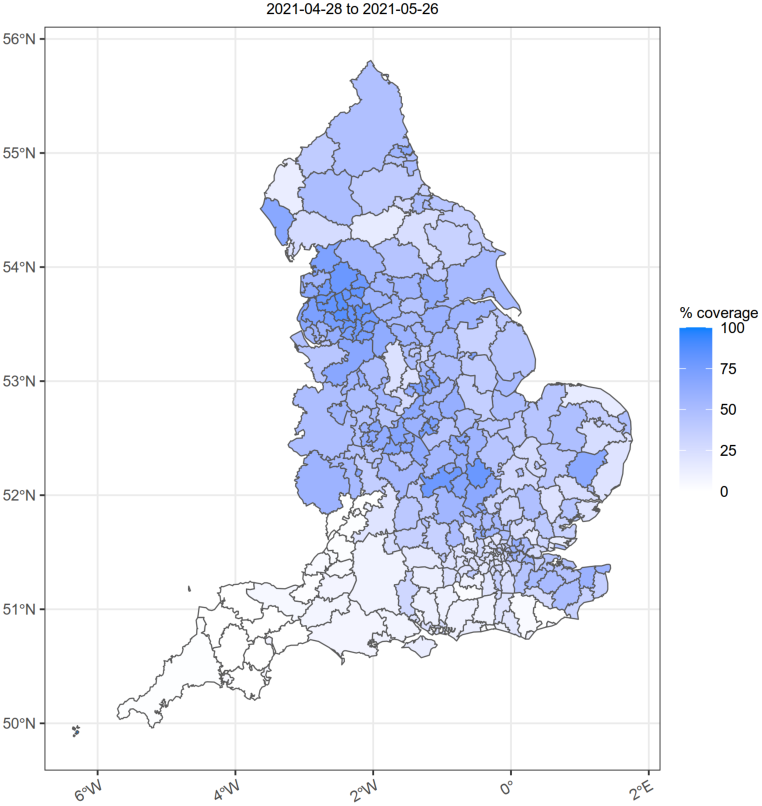
TaqPath test coverage in Pillar 2 by LTLA between 2021-04-17 and 2021-05-15.

#### 7. Epidemic curves

**Figure S7.1.**
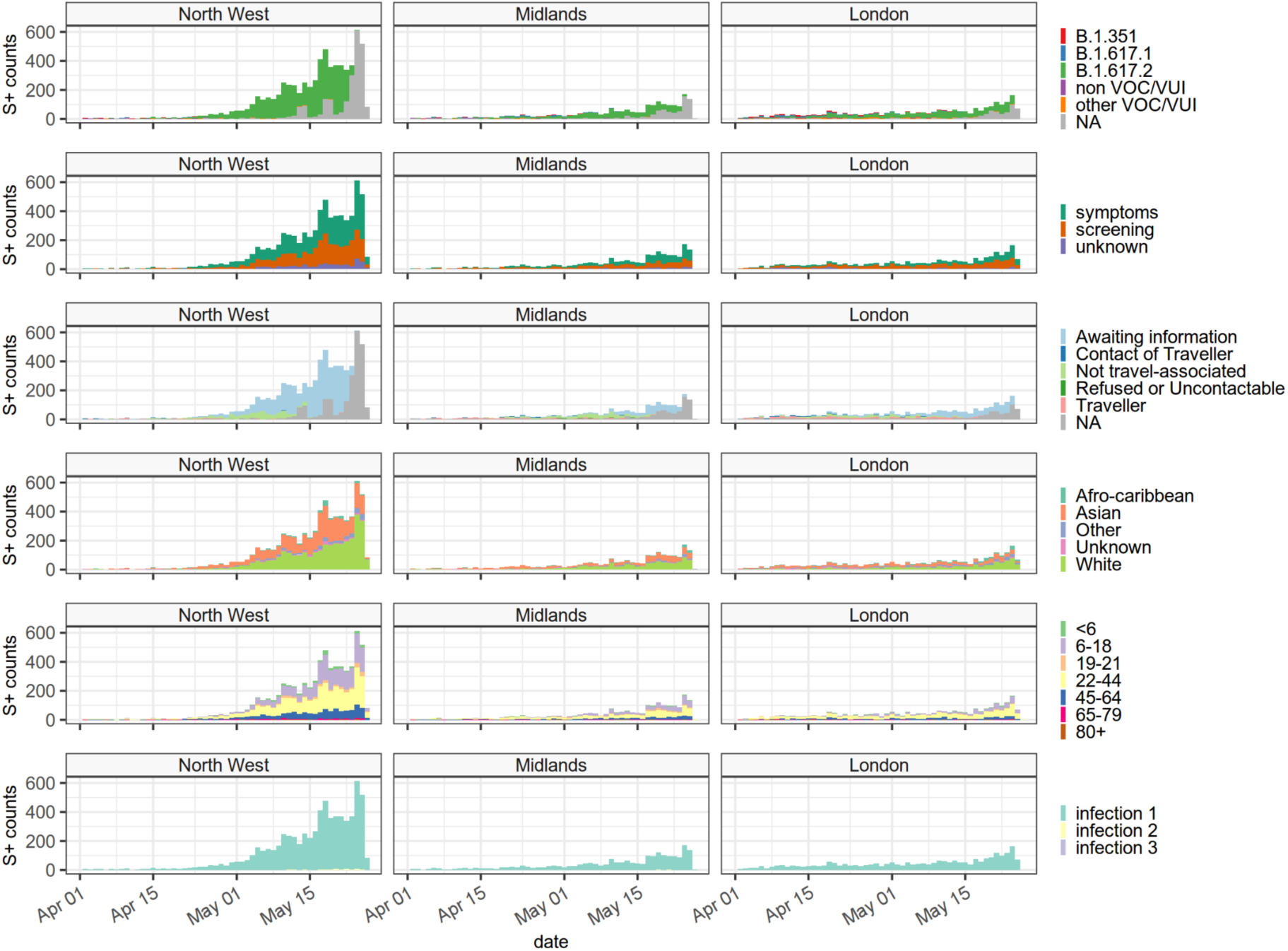
Epidemic curves for S-gene positive cases in the 3 NHSER regions under investigation. The NHS regions selected are those with the most reliable TaqPath coverage. The majority of cases to date have been in the North West. Cases are heterogeneous in age and ethnicity. At least half of all cases arise from symptomatic infection. Cases in grey are S-gene positive cases for which we have no sequencing information yet. The last 4 days of case counts will be revised upwards due to reporting delays, and are excluded from growth rate calculations.

**Figure S7.2.**
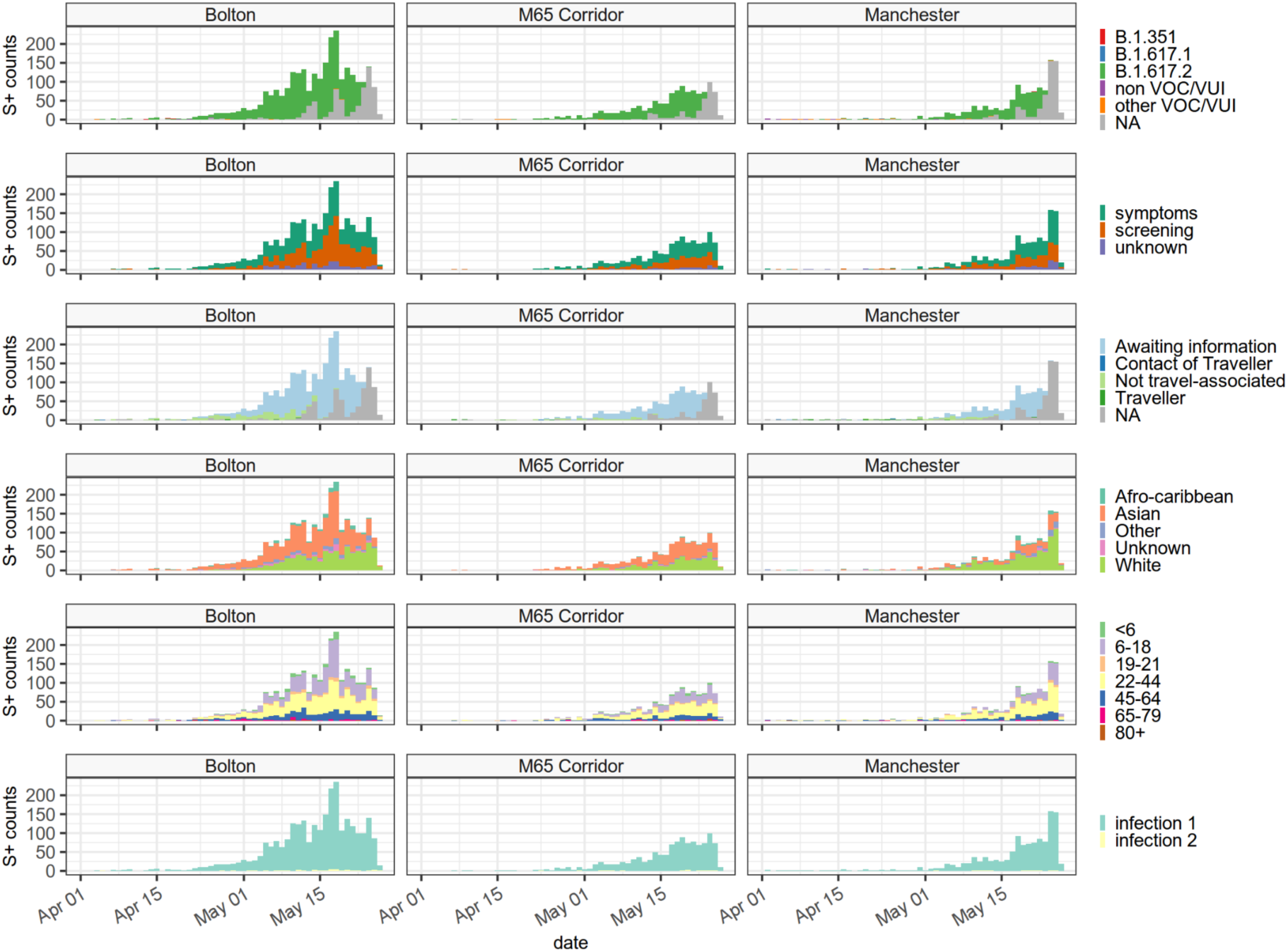
Epidemic curves for S-gene positive cases in the 3 smaller regions under investigation in the North West. As the first and most significantly affected region Bolton has had intensive case finding and testing efforts. Cases are seen to be reaching a plateau here. A similar picture is seen in the M65 Corridor. Case numbers in Manchester are still growing. The last 4 days of case counts will be revised upwards due to reporting delays, and are excluded from growth rate calculations.

**Figure S7.4.**
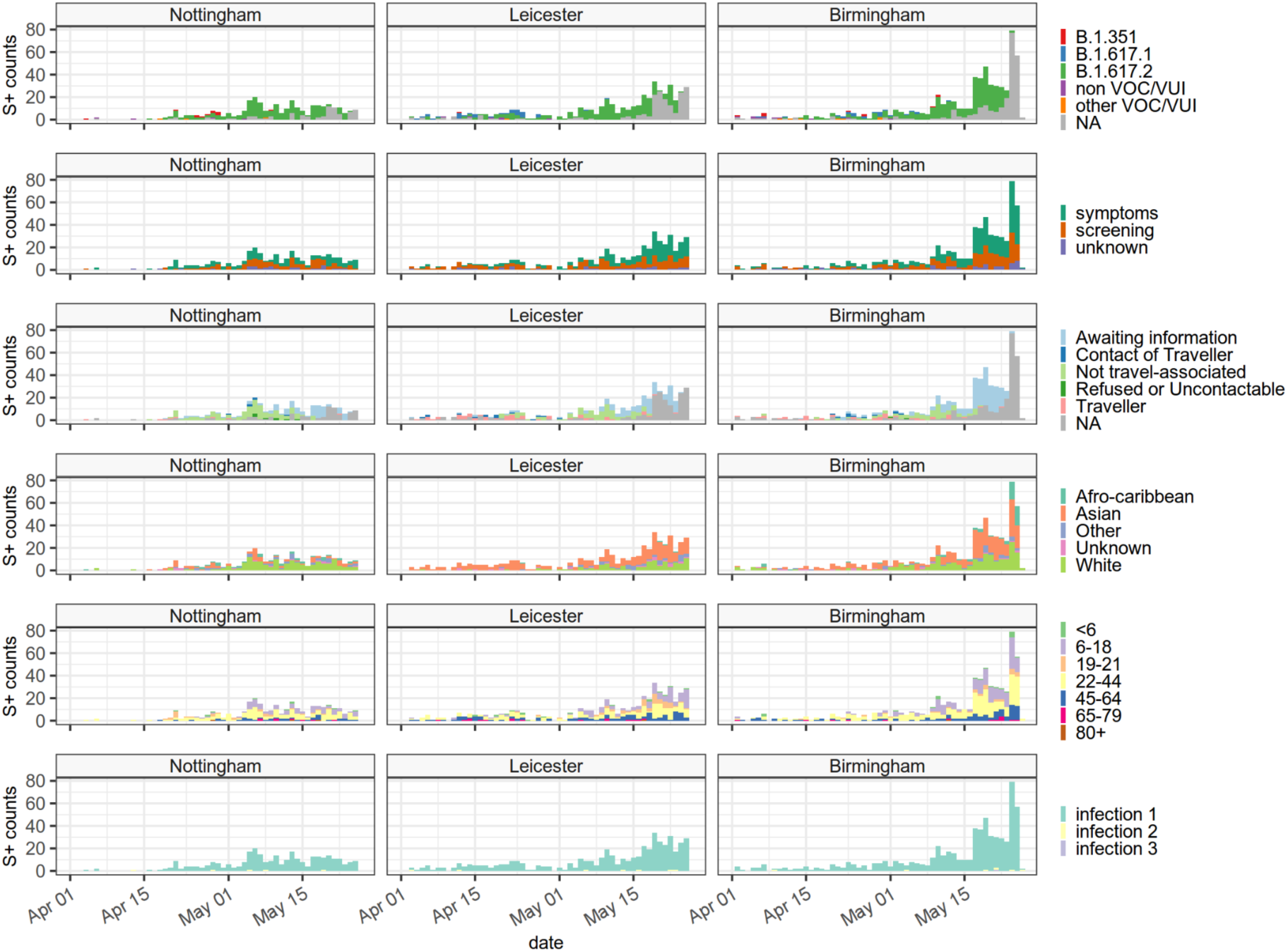
Epidemic curves for S-gene positive cases in the 3 smaller regions under investigation in the Midlands. Nottingham and Leicester had earlier imports of a variety of S-gene positive variants including B.1.351. More recently the sequencing has been dominated by B.1.617.2. Case growth in Nottingham has thus far been relatively slow, compared to the other two regions. Cases in Birmingham are increasing rapidly. The last 4 days of case counts will be revised upwards due to reporting delays, and are excluded from growth rate calculations.

